# Baseline Inflammatory Profiles in Moderate-to-Severe Depression and Differential Response to Intermittent Theta-Burst Stimulation

**DOI:** 10.1101/2025.07.16.25331663

**Authors:** Bruno Pedraz-Petrozzi, Jonas Wilkening, Nicole Ziegler, Martijn Arns, Alexander Sartorius, Roberto Goya-Maldonado

## Abstract

Major depressive disorder (MDD) is a biologically heterogeneous condition, with a subset of patients exhibiting elevated inflammatory markers associated with greater symptom burden and lower responsiveness to antidepressants. Treatment outcomes with repetitive transcranial magnetic stimulation (rTMS) are also highly variable, and inflammation may impair neuroplasticity, thereby modulating treatment efficacy. This exploratory study investigated whether baseline inflammatory protein signatures could influence the response to intermittent theta burst stimulation (iTBS). Using the Olink® Target 96 Inflammation panel, baseline inflammatory protein levels were analyzed in 54 patients with moderate-to-severe depression (baseline MADRS ≥20) undergoing iTBS treatment. K-means clustering identified two subgroups: Cluster 1 (n = 28) and Cluster 2 (n = 26). Cluster 2 showed the highest proportion of responders, whereas Cluster 1 showed the lowest. The clusters did not differ significantly in age, sex, medication status, randomization arm, or body mass index. Differential protein expression was assessed using volcano plots with false discovery rate (FDR) correction. Pathway enrichment analyses using Kyoto Encyclopedia of Genes and Genomes (KEGG) and Reactome databases identified biological processes associated with the clustering solution. Volcano analysis identified 10 differentially expressed proteins between clusters (FDR < 0.05 and |ΔNPX| ≥ 1). Enrichment analyses implicated cytokine–cytokine receptor interaction, chemokine signaling, and interleukin signaling pathways as characteristic of the lower-response subgroup. These findings indicate that baseline inflammatory signatures may help differentiate responsiveness to iTBS in moderate-to-severe depression, supporting future pathway investigations as a potential strategy for personalized psychiatry.

## 1. INTRODUCTION

Major depressive disorder (MDD) is a highly prevalent psychiatric condition and a leading cause of disability worldwide (World Health Organization, 2017). Diagnostic frameworks such as the DSM-5 define depression based on symptom severity, reflecting its association with functional impairment and healthcare burden (Beiser et al., 2019; Chen and Lin, 2011; Voelker et al., 2021; Zimmerman et al., 2018). However, growing evidence underscores the marked biological heterogeneity underlying this syndrome (Dinga et al., 2019; Drysdale et al., 2017).

In particular, a distinct immuno-inflammatory subtype of MDD has been described (Raison and Miller, 2011; Schmidt et al., 2016; Strawbridge et al., 2019). This subtype is characterized by low-grade systemic inflammation and elevated levels of C-reactive protein (CRP) and pro-inflammatory cytokines (e.g., IL-6, IL-1β, TNF-α), which have been associated with greater disorder severity (Gavril et al., 2024; Jeenger et al., 2017; Köhler-Forsberg et al., 2017; Orsolini et al., 2022; Osimo et al., 2019; Paganin and Signorini, 2024) and poorer treatment outcomes (Benedetti et al., 2021; Breit et al., 2023; Strawbridge et al., 2015), especially to antidepressants (Dahl et al., 2014; Liu et al., 2020; Uher et al., 2014; Yang et al., 2019).

Next to conventional pharmacotherapy and the gold-standard electroconvulsive therapy (ECT), repetitive transcranial magnetic stimulation (rTMS) has emerged as an effective, non-invasive neuromodulation treatment for patients with MDD, offering fewer side effects and improved tolerability. Among its protocols, intermittent theta burst stimulation (iTBS) has gained prominence due to its shorter session duration and comparable efficacy to conventional 10 Hz stimulation applied to the left dorsolateral prefrontal cortex (DLPFC) (Blumberger et al., 2018; Cole et al., 2022; Lee et al., 2023; Neuteboom et al., 2023). Nevertheless, response to iTBS remains highly variable (Kishi et al., 2024), underscoring the need for biomarkers that can predict and optimize treatment outcomes (Breit et al., 2023). Neuroinflammatory processes are increasingly recognized as modulators of synaptic plasticity and cortical excitability (Guo et al., 2023). Synaptic plasticity refers to the brain’s ability to strengthen or weaken synaptic connections, whereas cortical excitability refers to the responsiveness of neurons to stimulation (Guo et al., 2023; Lenz et al., 2021). These effects are thought to involve activation of inflammatory signaling cascades such as the Janus kinase/signal transducer and activator of transcription (JAK/STAT) and nuclear factor kappa-light-chain-enhancer of activated B cells (NF-κB) pathways. These pathways regulate cytokine signaling, glial cell reactivity, and neuroplastic processes implicated in depression (Feng et al., 2024; Kennedy and Silver, 2016; Miller and Raison, 2016). This has led to the hypothesis that peripheral inflammatory activity may influence the effectiveness of iTBS treatment. However, most neuromodulation studies have examined a narrow subset of cytokines, often yielding inconsistent results (Pedraz-Petrozzi et al., 2024). A more comprehensive characterization of inflammatory profiles at the systems level (i.e., considering multiple interacting pathways) may therefore clarify these inconsistencies and enable more precise, biomarker-guided stratification of treatment responses.

Recent advances in high-throughput proteomic technologies, such as the Olink platform, enable the simultaneous quantification of over 90 immune-related proteins with high sensitivity and reproducibility from small plasma volumes (Assarsson et al., 2014). These approaches enable the characterization of complex, pathway-level immune signatures, moving beyond single-marker associations toward biologically grounded ‘immune biotypes’ - that is, distinct patient subgroups defined by shared patterns of immune activity. By providing a systems-level view of inflammatory signaling, proteomic profiling can identify patient subgroups with underlying biological differences that are not apparent from conventional clinical classifications (Fillman et al., 2014; Mesleh et al., 2021; Steiner et al., 2017; Yang et al., 2023).

In this exploratory study, we used targeted inflammatory proteomics to determine whether distinct baseline immune signatures could distinguish subgroups of patients with moderate-to-severe depression (defined by baseline MADRS ≥20) undergoing iTBS based on treatment response. By applying data-driven clustering to the Olink proteomic data, we aimed to identify inflammatory profiles based on biological markers, thereby better understanding the mechanisms underlying variability in neuromodulation outcomes. This approach aligns with the broader goal of precision psychiatry, which seeks to move beyond symptom-based classification toward biologically informed patient stratification.

## 2. MATERIALS AND METHODS

### 2.1. Study design

This exploratory analysis draws on data from a larger clinical trial that investigated the imaging, psychometric, and neuromodulatory effects of iTBS in patients with MDD (Pre-mapping Networks for Brain Stimulation 2; clinicaltrials.gov/show/NCT05260086). The main study followed a six-week longitudinal design, which has been described previously (Wilkening et al., 2022).

The trial used a quadruple-blinded, randomized, sham-controlled, crossover design, ensuring that participants, care providers, investigators, and outcome raters were unaware of treatment allocation. Each participant completed two one-week stimulation phases (one with active iTBS and one with sham stimulation). The order of these phases (active-sham or sham-active) was randomized for each participant using pre-coded sequences generated in MATLAB (The MathWorks, Inc., Natick, MA, USA). The principal investigator supervised the study while maintaining blinding from all participant interactions (Wilkening et al., 2022; Wilkening and Goya-Maldonado, 2025).

Additionally, a parallel pseudorandomized design was implemented to assess the clinical utility of personalized stimulation targeting. Participants either received stimulation at the standard F3 scalp location (following the international 10–20 EEG system) or at an individually selected site within the left DLPFC, identified via resting-state functional MRI. For this analysis, we did not differentiate between fixed and individualized targeting, as both approaches employed similar methods (Wilkening et al., 2022; Wilkening and Goya-Maldonado, 2025). Previous analyses of this cohort found no significant difference in treatment outcomes based on stimulation target group (Wilkening et al., 2025a); therefore, the entire sample was combined for the present study.

Depressive symptoms were evaluated using the Montgomery-Åsberg Depression Rating Scale (MADRS), administered by trained clinicians at baseline and again at the end of each week, allowing for the assessment of symptom changes over the six-week study period (Wilkening et al., 2022; Wilkening and Goya-Maldonado, 2025).

### 2.2. Participants

The study protocol complied with the Declaration of Helsinki and received approval from the Ethics Committee at the University Medical Center Göttingen (UMG). Before taking part, all participants provided both verbal and written informed consent after receiving thorough information about the study procedures (Wilkening and Goya-Maldonado, 2025). Participants were eligible if they were adults aged 18 to 60 years, met DSM-5 criteria for major depressive disorder (MDD), and were experiencing a moderate-to-severe depressive episode. Only those with a baseline MADRS score of 20 or higher were included in this exploratory study.

Trained psychiatrists confirmed MDD diagnoses using the Structured Clinical Interview for DSM-5 Disorders, Clinical Version (SCID-5-CV), and determined depression severity with the MADRS (Hengartner et al., 2020; Montgomery and Åsberg, 1979; Wilkening and Goya-Maldonado, 2025). Exclusion criteria covered any conditions that would contraindicate rTMS, such as a history of epilepsy, significant neurological disorders, pregnancy, or the presence of metallic implants (Wilkening and Goya-Maldonado, 2025).

Participants with evidence of acute or systemic inflammation based on baseline C-reactive protein (CRP) or white blood cell count (WBC) were excluded. Specifically, individuals were excluded if CRP exceeded 10 mg/L or if the WBC was greater than 11,000 cells/μL, as these thresholds are commonly used to indicate more overt acute inflammatory or infectious processes (Gabay and Kushner, 1999; Haburchak and Alchreiki, 2020; Mank et al., 2025; Myers et al., 2004; Nehring et al., 2025). To maintain consistency, participants who used pharmacotherapy were required not to have any changes for at least two weeks prior to the study enrollment and throughout its duration. Concomitant medication was categorized for analysis as no medication at all, non-antidepressant medication, other antidepressants (non-SSRIs), and SSRIs. Antidepressants represented in the sample included SSRIs (e.g., escitalopram, sertraline, and citalopram) and non-SSRI antidepressants (e.g., venlafaxine, bupropion, duloxetine, mirtazapine, agomelatine, trazodone, and moclobemide). Frequent monitoring of serum medication levels ensured treatment adherence, and these values were incorporated into the statistical analyses to control for confounding effects on clinical outcomes (Wilkening and Goya-Maldonado, 2025). Both inpatients and outpatients were included in the sample, but all neuromodulation sessions were conducted in an outpatient setting. Details regarding study procedures and the underlying rationale have been published previously (Wilkening et al., 2022; Wilkening and Goya-Maldonado, 2025).

### 2.3. iTBS treatment

The technical specifications and details of the iTBS protocol applied in this study are available in the published study protocol (Wilkening et al., 2022; Wilkening and Goya-Maldonado, 2025). Briefly, the iTBS was administered using a MagVenture system equipped with MagOption and a figure-of-eight MCF-B65 A/P cooled coil, operated by trained clinical staff. Stimulation consisted of four sessions per day, each lasting approximately 10 minutes, with at least 20-minute intervals between sessions in accordance with established practices to allow for inter-session spacing (Tse et al., 2018; Wilkening and Goya-Maldonado, 2025). The iTBS paradigm involved bursts of three pulses at 50 Hz delivered at 5 Hz, with 2-second trains and 8-second rest periods. Each session comprised 60 bursts (a total of 1,800 pulses per session), amounting to 7,200 pulses per day and a cumulative 36,000 pulses during the entire treatment period.

We determined the resting motor threshold (RMT) daily using electromyography from the right first dorsal interosseous muscle. RMT was defined as the lowest intensity that evoked motor potentials of ≥50 µV in at least 5 out of 10 consecutive trials. Stimulation intensity was set at 110% of the individual RMT. For sham stimulation, we rotated the coil 180 degrees to simulate the sound associated with active stimulation while reducing the actual effective output. To further mimic the somatosensory aspects, we applied transcutaneous electrical nerve stimulation (tENS) electrodes to the scalp during both real and sham conditions. To evaluate the integrity of the blinding procedure, participants rated their experience using a visual analogue scale (Wilkening and Goya-Maldonado, 2025).

### 2.4. Proteomics: Inflammatory Marker Assessment and Enrichment Analysis

Peripheral venous blood samples were collected at the start of the study in tubes containing ethylenediaminetetraacetic acid (EDTA). These initial samples were drawn before group assignment (whether active-sham or sham-active). Plasma was isolated by spinning the tubes at 3,000 rpm for 15 minutes and then stored at –80°C in the university’s research facilities until needed for analysis.

To measure markers of inflammation, we used the Olink® Target 96 Inflammation panel (Olink Proteomics, Uppsala, Sweden). This panel quantifies 92 inflammation-related proteins using proximity extension assay (PEA) technology. Each protein measurement is obtained using a specific pair of oligonucleotide-tagged antibodies. Once both antibodies bind to the target protein, the DNA strands are brought together, allowing them to hybridize and be amplified by real-time PCR. Data are presented as Normalized Protein Expression (NPX) values, which are log_2_-transformed relative units standardized by the manufacturer (Assarsson et al., 2014; Yang et al., 2023). All Olink assays took place at the Fraunhofer Institute for Translational Medicine and Pharmacology in Frankfurt am Main, Germany.

Before conducting the analyses, we assessed the quality of the NPX data. All 92 inflammatory proteins measured by the Olink® panel were present. The majority showed excellent detectability, with 82 of 92 proteins having more than 95% of NPX values above the limit of detection (LOD), and a median missingness of 0%. Overall missingness averaged ∼15%, but this was almost entirely attributable to 10 low-abundance proteins with high proportions of LOD-flagged values (β-NGF, IL-2, IL-2RB, IL-20, IL-13, IL-33, LIF, IL-1α, NRTN, TSLP). Importantly, in the Olink system, values below the LOD are still provided as NPX measurements and flagged with LOD indicators rather than being removed or coded as missing (Bevital AS, 2023; Pramod Shinde, 2021). Because these <LOD values remain in the dataset, no actual data loss occurs, and no exclusion based on missingness was required; all 92 proteins were therefore retained for analysis. We also did not apply any imputation procedures (e.g., LOD substitution, mean imputation, or regression-based estimation). Given the relatively small sample size, imputation could artificially reduce variability or introduce spurious associations. Consistent with recent recommendations suggesting that avoiding imputation can yield more robust results in smaller proteomic datasets (Bramer et al., 2021), all NPX values (including those flagged as <LOD) were analyzed as provided (Bevital AS, 2023; Pramod Shinde, 2021). This approach avoids distorting the natural biological variation that may occur when few inflammatory markers fall below the detection range in specific subgroups.

### 2.5. Statistical analysis

Initially, to identify potential subgroups based on baseline inflammatory marker expression, we conducted a k-means cluster analysis. Before clustering, all NPX values were standardized so that each of the 92 proteins would contribute equally, correcting for differences in scale. K-means clustering was performed on the standardized NPX values of all 92 inflammatory proteins from the Olink panel for each participant, such that each protein contributed equally after correction for differences in scale. No feature selection was performed before clustering. The Hartigan–Wong algorithm was employed for clustering, with a maximum of 1,000 iterations to ensure convergence.

To determine the optimal number of clusters, we used the Elbow method, Gap Statistics, and the average silhouette width, all applied to the standardized biomarker data. These methods consistently indicated that two clusters were optimal (see **Supplementary Material 1**). **Figure 1a** provides a graphical representation of the distributions of inflammatory markers by cluster.

**Figure 1.**
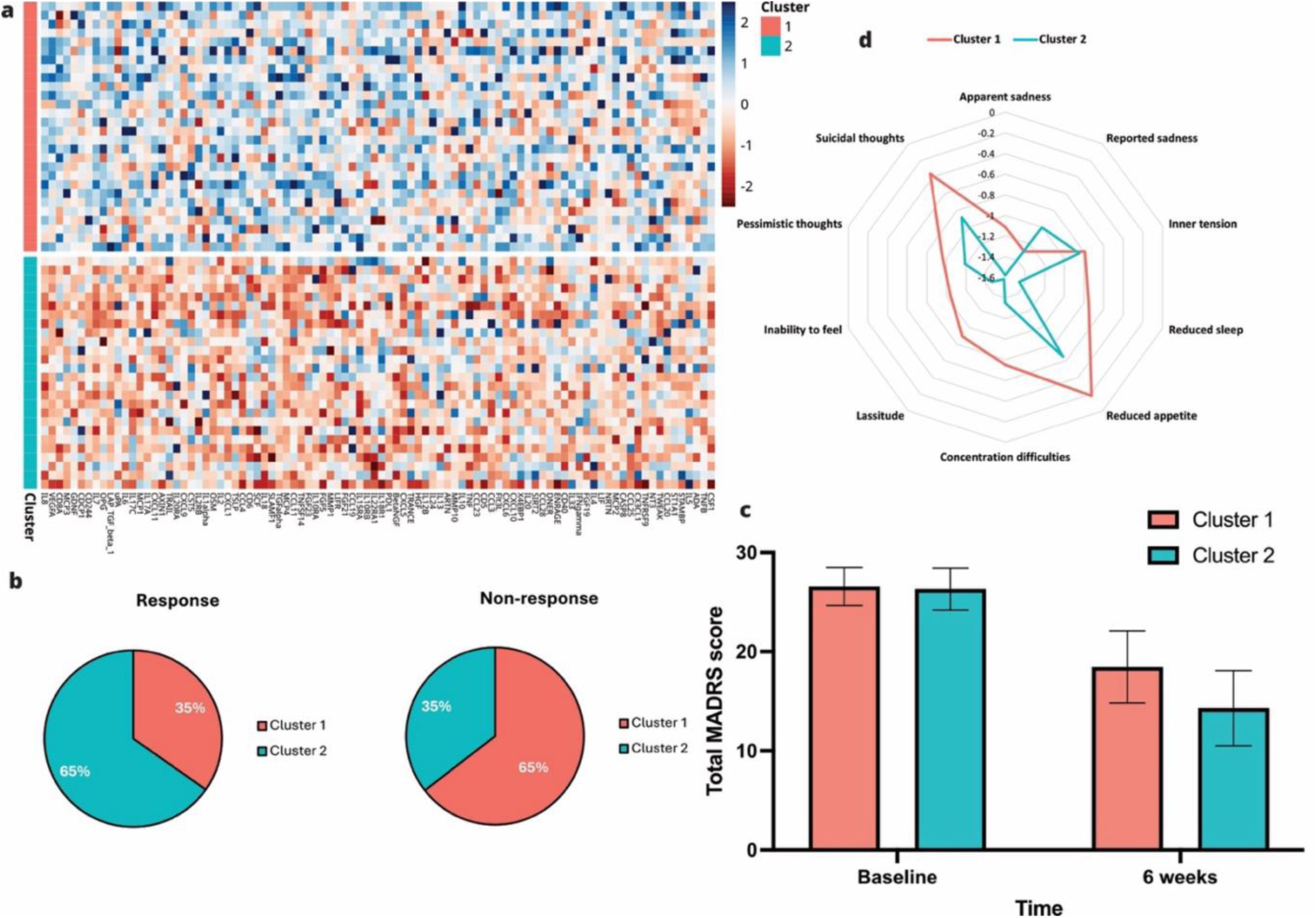
(a–d). Cluster profiles, symptom-level and response-related differences between clusters. **(a)** Cluster profiles based on standardized inflammatory marker expression. The y-axis represents the cluster profiles, and the x-axis shows the z-scores of the 92 inflammatory proteins included in the Olink panel in NPX. The plot illustrates the distinct inflammatory profiles of each cluster derived from k-means clustering. **(b)** Pie charts showing the proportion of responders and non-responders in each cluster (response defined as ≥50% reduction in MADRS total score); Cluster 2 includes a higher percentage of responders, whereas Cluster 1 includes a higher percentage of non-responders. **(c)** Bar plot showing cluster-wise mean change in MADRS total score with 95% CIs, illustrating broadly similar overall improvement across clusters. **(d)** Descriptive radar plot of the mean change (Δ) in individual MADRS item scores from baseline to post-treatment for each cluster. Negative values indicate symptom improvement. Item-level MADRS changes are shown for descriptive purposes only and were not formally compared statistically.

We evaluated the stability and robustness of the two-cluster solution using bootstrap resampling (1,000 iterations) with the clusterboot() function (Fehrer et al., 2025; Hennig, 2024, 2007). In this analysis, the clustering algorithm was repeatedly applied to bootstrap samples of the dataset, and the cluster consistency across resamples was assessed using the Jaccard similarity coefficient. According to established benchmarks (Fehrer et al., 2025; Hennig, 2024, 2007), Jaccard values above 0.85 indicate high stability, while values below 0.50 signal cluster dissolution (Fehrer et al., 2025; Hennig, 2024, 2007). The two clusters yielded Jaccard indices of 0.955 and 0.951, showing high stability and reproducibility; none were dissolved. By contrast, cluster configurations with k = 5 or k = 10 generated fragmented clusters with lower stability (multiple Jaccard < 0.50). These findings indicate that a two-cluster solution captures the most reproducible and biologically relevant heterogeneity in our study population while minimizing the risk of overfitting.

After defining the cluster structure, we characterized the resulting groups in terms of demographic (age, sex), anthropometric (body mass index), and treatment-related variables (medication class, treatment resistance, randomization arm), as well as change in depressive symptoms. Changes in the MADRS total score (ΔMADRS = post-treatment MADRS minus baseline MADRS) were compared between clusters using t-tests. This allowed examination of potential differences in treatment outcomes between the subgroups. Categorical variables, such as sex, medication class, baseline treatment resistance, randomization group, and responder rates (defined as ≥ 50% reduction in MADRS total score), were analyzed by chi-square tests. Symptom-level differences within the two-cluster solution were explored descriptively with radar plots showing changes in individual MADRS items from baseline to post-treatment across clusters. To assess whether concomitant medication might confound the association between inflammatory cluster membership and treatment response, we performed sensitivity analyses including medication category, medication-free status, and baseline medication load. Concerning the latter, to account for the psychotropic medication load, following our previous publication (Wilkening et al., 2025b), we summed for each individual the estimated intake dose of each substance at baseline, based on the interview or, when possible, on the circulating blood levels (below = 1, normal = 2, or above = 3 of the expected doses for each medication). Because some medication subgroups were small, adjusted responder models were estimated using Firth penalized logistic regression.

We next performed differential expression analyses for the inflammatory proteins. For each protein, we calculated mean NPX differences between clusters and used Welch’s t-tests for statistical comparison. Volcano plots display the relationship between log₂ NPX differences and significance. To correct for multiple testing, we applied the Benjamini-Hochberg FDR method. Proteins were considered significantly differentially expressed if |ΔNPX| > 1 (about a two-fold change on the log₂ scale) and Benjamini-Hochberg adjusted p-value < 0.05, in line with recent recommendations in Olink-based studies (Fredolini et al., 2024; Tayal et al., 2022; Wen et al., 2025). This threshold exceeds expected technical variation, helping to prioritize robust, biologically meaningful effects. Standardized effect sizes, such as Cohen’s d, Hedges’ g (for small sample correction), and Glass’s Δ (for variance differences), were calculated for each significant protein to quantify between-group differences.

In the final step, we analyzed pathways to provide a biological context for our results. Proteins exhibiting significant expression differences between clusters were subjected to pathway enrichment analysis using the Kyoto Encyclopedia of Genes and Genomes (KEGG) and Reactome databases. This allowed us to identify biological and signaling processes associated with proteins whose expression varied between clusters. Enrichment results are presented with q-values and false discovery rates (FDR), with statistical significance defined as q < 0.05 and FDR < 0.05. All statistics and visualizations, including enrichment analyses, were performed in RStudio (version 2024.12.1+563).

## 3. RESULTS

### 3.1. Subgroup characterization: general characteristics, clinical outcomes and symptom interaction

The sample comprised 54 participants, with 28 assigned to Cluster 1 and 26 to Cluster 2 (**Table 1**). The two clusters did not differ significantly in demographic or baseline clinical characteristics, including age, sex, BMI, treatment resistance, medication class, or randomization order (all p > 0.12; **Table 1**). With regard to treatment outcome, Cluster 2 showed a significantly higher proportion of responders than Cluster 1 (p = 0.031; **Table 1**; **Figure 1b**). By contrast, baseline MADRS scores and mean change in total MADRS score did not differ significantly between clusters (**Table 1**; **Figure 1c**). Thus, although the mean reduction in total MADRS score was comparable across groups, a larger proportion of patients in Cluster 2 achieved clinically meaningful symptom relief, defined as a reduction of at least 50% in MADRS total score. This suggests that baseline inflammatory differences may be more closely related to the likelihood of achieving a categorical response than to average symptom change.

**Table 1.**
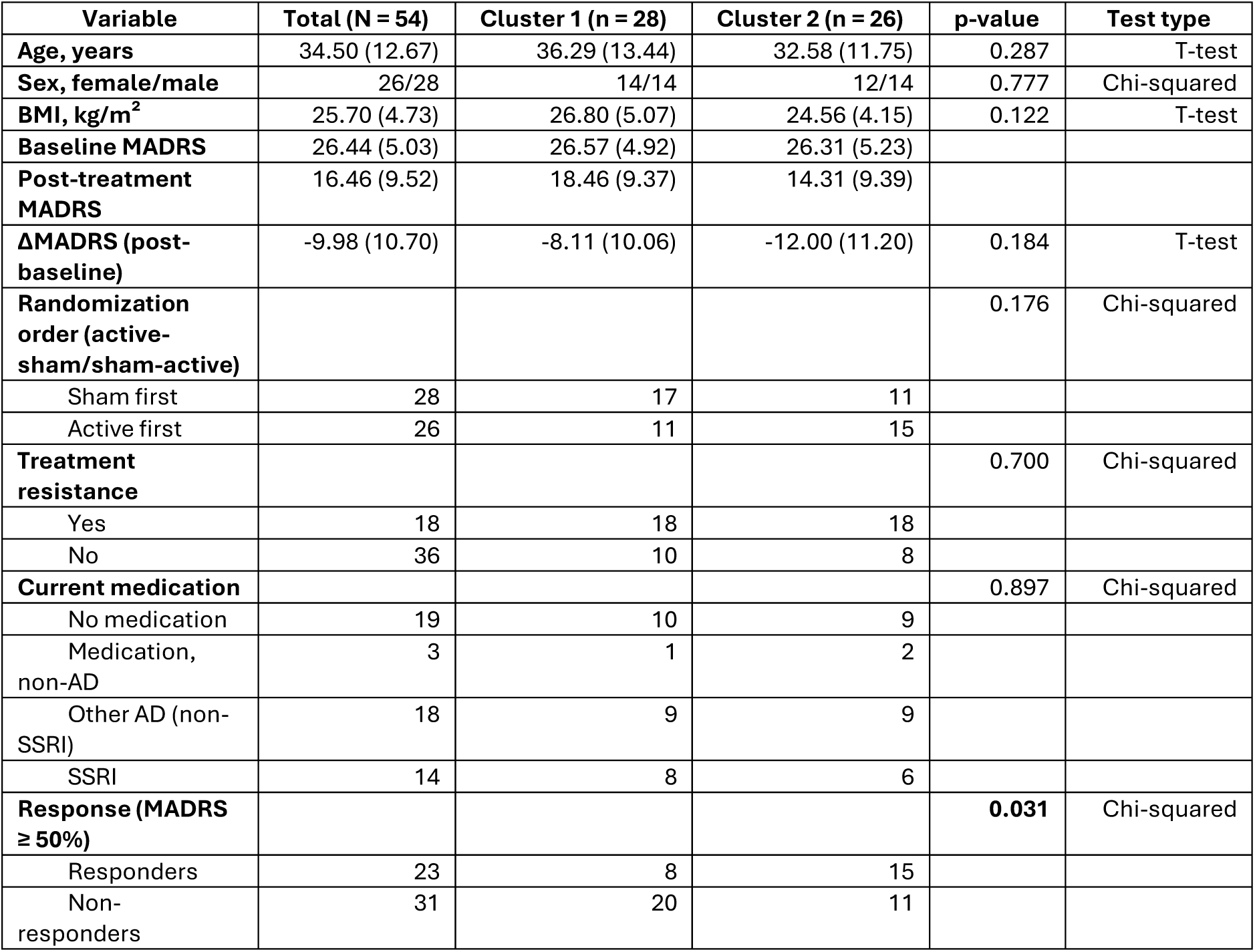
General and clinical characteristics of the two-cluster solution (derived from baseline inflammatory profiles), including statistical comparisons across groups. Data are presented as mean (SD) or counts. Group differences were assessed using T-tests or chi-squared tests. Response was defined as ≥50% reduction in MADRS score from baseline. Marked in bold are p-values less than or equal to 0.05. Abbreviations: BMI = body mass index, AD = antidepressants; MADRS = Montgomery–Åsberg Depression Rating Scale; SSRI = selective serotonin reuptake inhibitors.

| Variable | Total (N = 54) | Cluster 1 (n = 28) | Cluster 2 (n = 26) | p-value | Test type |
| --- | --- | --- | --- | --- | --- |
| Age, years | 34.50 (12.67) | 36.29 (13.44) | 32.58 (11.75) | 0.287 | T-test |
| Sex, female/male | 26/28 | 14/14 | 12/14 | 0.777 | Chi-squared |
| BMI, kg/m<sup>2</sup> | 25.70 (4.73) | 26.80 (5.07) | 24.56 (4.15) | 0.122 | T-test |
| Baseline MADRS | 26.44 (5.03) | 26.57 (4.92) | 26.31 (5.23) |  |  |
| Post-treatment MADRS | 16.46 (9.52) | 18.46 (9.37) | 14.31 (9.39) |  |  |
| ΔMADRS (post-baseline) | -9.98 (10.70) | -8.11 (10.06) | -12.00 (11.20) | 0.184 | T-test |
| Randomization order (active-sham/sham-active) |  |  |  | 0.176 | Chi-squared |
| Sham first | 28 | 17 | 11 |  |  |
| Active first | 26 | 11 | 15 |  |  |
| Treatment resistance |  |  |  | 0.700 | Chi-squared |
| Yes | 18 | 18 | 18 |  |  |
| No | 36 | 10 | 8 |  |  |
| Current medication |  |  |  | 0.897 | Chi-squared |
| No medication | 19 | 10 | 9 |  |  |
| Medication, non-AD | 3 | 1 | 2 |  |  |
| Other AD (non-SSRI) | 18 | 9 | 9 |  |  |
| SSRI | 14 | 8 | 6 |  |  |
| Response (MADRS $\geq 50\%$) | | | | **0.031** | Chi-squared |
| Responders | 23 | 8 | 15 |  |  |
| Non-responders | 31 | 20 | 11 |  |  |

Exploratory analysis of symptom-level changes (ΔMADRS items) revealed broadly similar improvement patterns across clusters, with reductions observed across all MADRS items (**Table 1**; **Figure 1d**). Descriptively, Cluster 2 showed greater improvement across several symptom domains, particularly reduced sleep, lassitude, inability to feel, and suicidal thoughts. However, these item-level differences were exploratory and descriptive, were not formally tested statistically, and should therefore be interpreted with caution.

### 3.2. Sensitivity analysis for medication-related confounding

To assess whether medication use alone might explain the association between inflammatory cluster membership and treatment response, we performed additional sensitivity analyses. Medication category was not significantly associated with cluster assignment (Chi-square test: χ²(3) = 0.60, p = 0.897) or responder status (Chi-square test: χ²(3) = 3.11, p = 0.375). Likewise, baseline medication load did not differ significantly by cluster or responder status (all p > 0.59). Because some medication subgroups were small, adjusted responder models were estimated using Firth penalized logistic regression. In these analyses, cluster membership remained significantly associated with treatment response after adjustment for medication category (OR = 3.64, 95% CI 1.21–11.83, p = 0.021), baseline medication load (OR = 3.19, 95% CI 1.09–9.90, p = 0.034), and simplified medication status (medication-free vs. medicated; OR = 3.30, 95% CI 1.11–10.48, p = 0.031). Full results are provided in Supplementary Material 1.

### 3.3. Inflammatory parameters - differences between Cluster 1 and 2

After identifying significant differences in symptom alleviation between Clusters 1 and 2, we examined their inflammatory marker profiles, as these clusters represented opposite extremes: Cluster 2 included the highest proportion of treatment responders, whereas Cluster 1 contained the fewest.

To visualize and quantify molecular differences, we generated a volcano plot based on NPX values, with p-values adjusted for multiple comparisons using the Benjamini–Hochberg FDR procedure. Markers were considered significant when both criteria were met: FDR < 0.05 and an absolute NPX difference ≥ 1 (log₂ scale). Of the 92 inflammatory proteins analyzed, 47 showed FDR-adjusted p < 0.05 (including MCP-4, CXCL6, CXCL5, IL-8, IL-7, VEGFA, CXCL1, MCP-1, CXCL11, OSM, MMP-1, HGF, CCL4, LAP TGF-β1, CD40, β-NGF, TNFSF14, CCL11, CSF1, MCP-2, LIFR, CCL28, FGF5, IL-2, AXIN1, TWEAK, TGF-α, FGF23, IL-6, MCP-3, CXCL10, IL-18, CD244, TSLP, CD8A, CCL3, IL10RB, IL20, LIF, SCF, IL15RA, CX3CL1, TNF, OPG, IL24, CCL23, and CCL25; **Supplementary Material 2**).

However, only ten proteins satisfied both significance thresholds (FDR < 0.05 and |ΔNPX| ≥ 1), specifically MCP-4, CXCL6, IL-7, TNFSF14, CXCL1, OSM, CXCL5, IL-8, CXCL11, and MMP-1. We applied the additional |ΔNPX| ≥ 1 threshold to prioritize proteins that were not only statistically significant, but also associated with larger effect sizes. This criterion further emphasized differences likely to be biologically meaningful and greater than the expected technical variability of the assay. These outcomes are shown in the volcano plot (**Figure 2**) and the corresponding boxplots (**Figure 3**), which illustrate the expression differences between Clusters 1 and 2.

**Figure 2.**
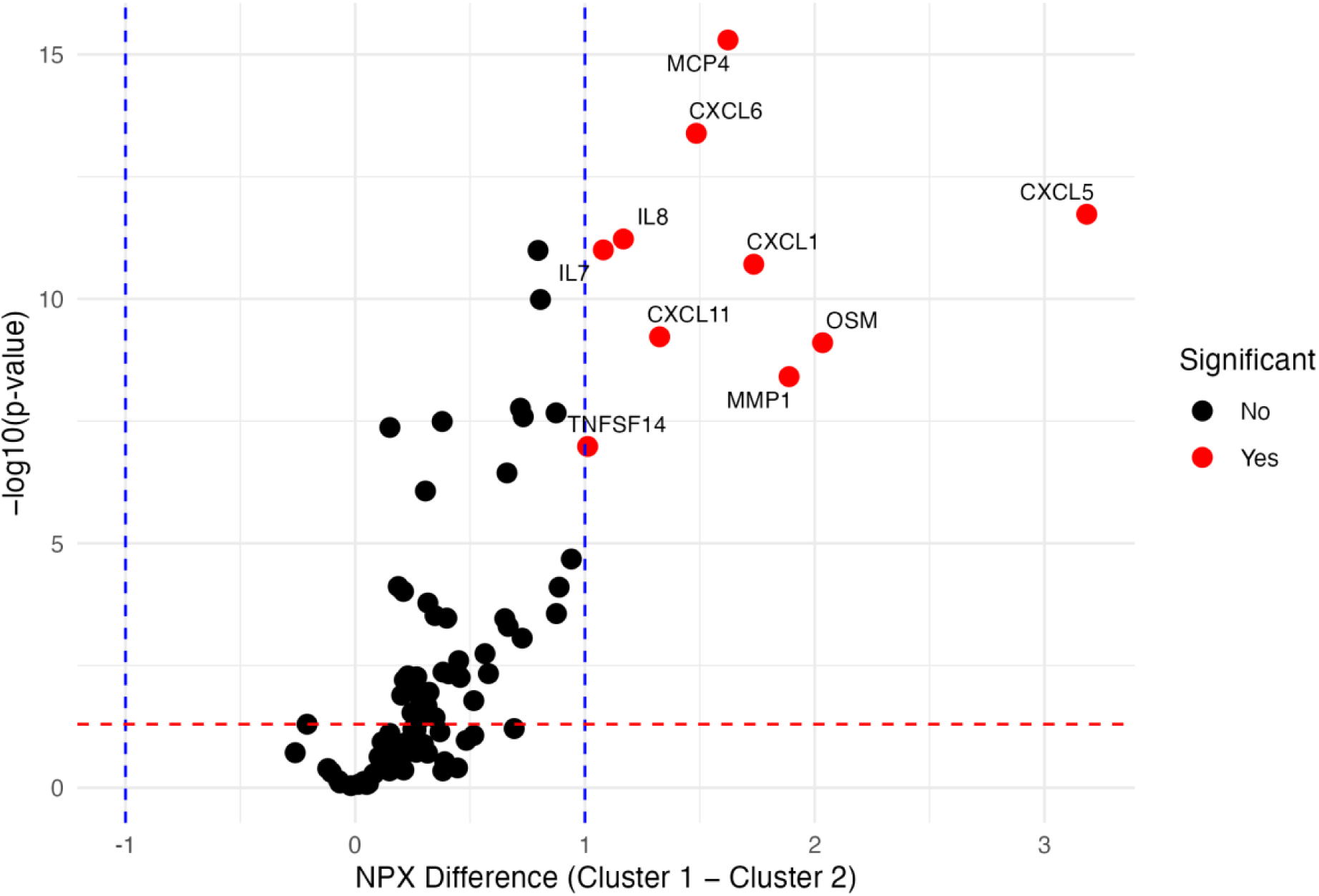
Volcano plot of NPX differences between Cluster 1 and Cluster 2. Vertical blue dotted lines denote NPX differences of +1 and –1. The horizontal red dashed line indicates the FDR-adjusted p-value threshold of 0.05. Abbreviations: NPX = Normalized Protein Expression; IL-7 = Interleukin-7; IL-8 = Interleukin-8; CXCL11 = C-X-C motif chemokine ligand 11; CXCL1 = C-X-C motif chemokine ligand 1; OSM = Oncostatin M; MMP-1 = Matrix metalloproteinase-1; CXCL6 = C-X-C motif chemokine ligand 6; MCP-4 = Monocyte chemotactic protein-4; CXCL5 = C-X-C motif chemokine ligand 5; TNFSF14 = TNF Superfamily Member 14.

**Figure 3.**
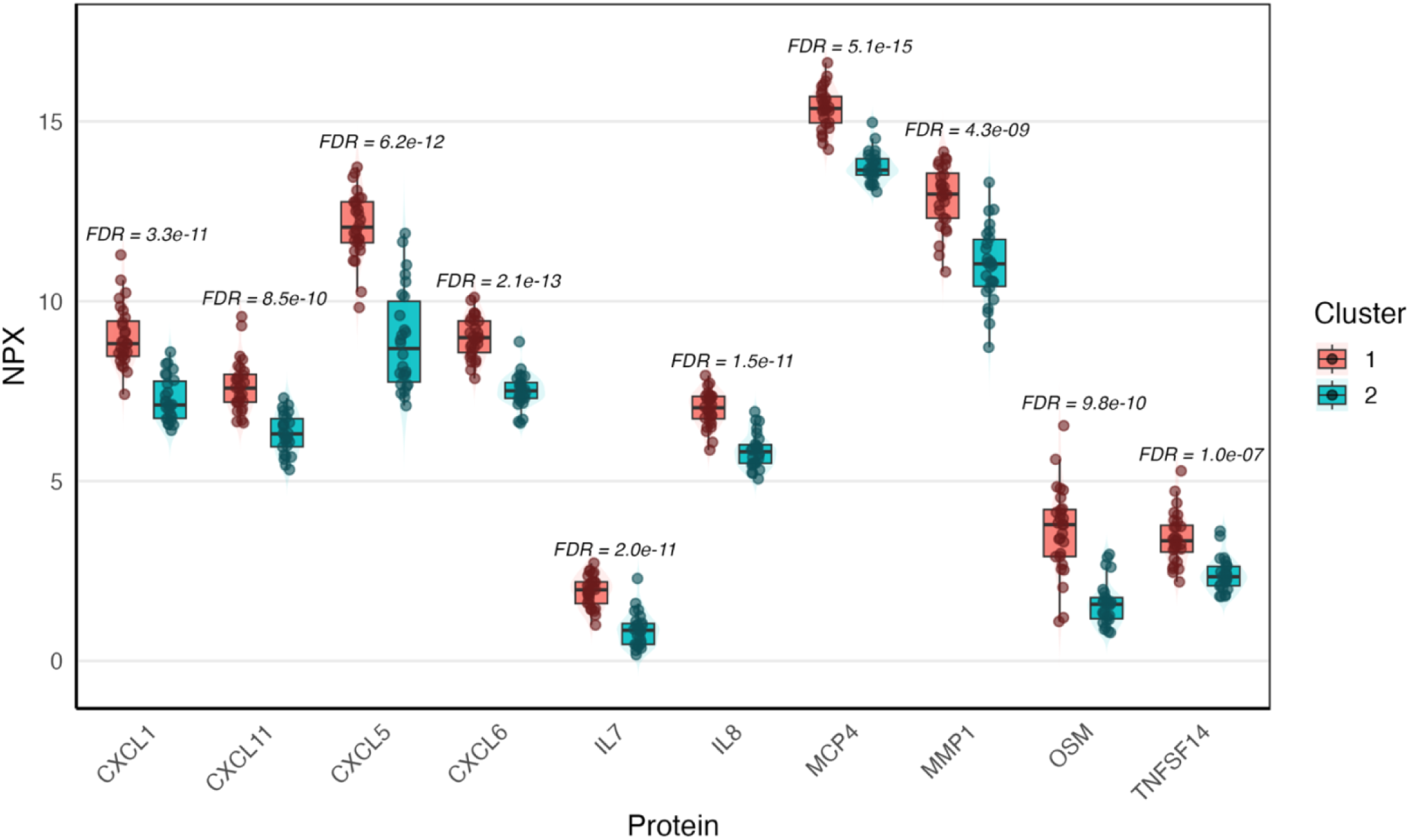
Boxplots of baseline NPX values for ten significantly different inflammatory proteins between Cluster 1 and Cluster 2. Clusters correspond to patient subgroups with the highest (Cluster 2) versus the lowest (Cluster 1) iTBS response rates; FDR-adjusted p-values are indicated above each plot. Abbreviations: NPX = Normalized Protein Expression; IL-7 = Interleukin-7; IL-8 = Interleukin-8; CXCL11 = C-X-C motif chemokine ligand 11; CXCL1 = C-X-C motif chemokine ligand 1; OSM = Oncostatin M; MMP-1 = Matrix metalloproteinase-1; CXCL6 = C-X-C motif chemokine ligand 6; MCP-4 = Monocyte chemotactic protein-4; CXCL5 = C-X-C motif chemokine ligand 5; TNFSF14 = TNF Superfamily Member 14.

The |ΔNPX| ≥ 1.0 criterion ensured that only proteins showing at least a two-fold relative difference between clusters were included, emphasizing changes that are likely to be biologically relevant and greater than the expected technical variability of the assay. To provide a more comprehensive assessment of these differences, p-values were supplemented with standardized effect-size metrics for all pairwise cluster comparisons, with particular attention to Clusters 1 and 2, where the greatest disparities were observed. In these analyses, Cohen’s d (with 95% confidence intervals), Hedges’ g (corrected for small sample bias), and Glass’s Δ (adjusting for unequal variances) were computed for each relevant protein. Across these metrics, the key proteins differentiating the clusters demonstrated consistently large effect sizes (e.g., MCP4: Cohen’s d = 3.18, Hedges’ g = 3.13; IL7: Cohen’s d = 2.40, Hedges’ g = 2.36; **Supplementary Material 1**). These results collectively reveal strong and biologically significant differences between clusters, even given the relatively small subgroup sizes. Furthermore, all proteins that met the significance criteria were quantifiable for all participants above the detection limit (see Methods).

### 3.4. Enrichment analysis

To explore the biological significance of the baseline differentially expressed proteins between clusters, we performed KEGG pathway enrichment analysis using the full inflammatory proteome as background. Several immune and inflammatory pathways were significantly enriched (**Figure 4a**). The most prominent pathway was cytokine–cytokine receptor interaction (FDR = 9.94 × 10⁻¹²; q = 5.37 × 10⁻¹²), highlighting broad involvement of intercellular signaling mechanisms that regulate immune activation. Additional enriched pathways included viral protein interaction with cytokine and cytokine receptor (FDR = 2.65 × 10⁻¹¹; q = 1.43 × 10⁻¹¹), chemokine signaling (FDR = 1.66 × 10⁻⁷; q = 8.98 × 10⁻⁸), and IL-17 signaling (FDR = 1.70 × 10⁻⁷; q = 9.19 × 10⁻⁸), all of which are closely associated with pro-inflammatory cytokine cascades. Pathways linked to rheumatoid arthritis (FDR = 1.70 × 10⁻⁷; q = 9.19 × 10⁻⁸) and NF-κB signaling (FDR = 1.83 × 10⁻⁵; q = 9.88 × 10⁻⁶) further indicate activation of canonical immune processes. Other relevant pathways, including TNF signaling, Toll-like receptor signaling, and JAK–STAT signaling, were also enriched, reinforcing the role of inflammatory signal transduction in these proteomic differences. A complete list of enriched KEGG pathways is provided in **Supplementary Material 1**.

**Figure 4.**
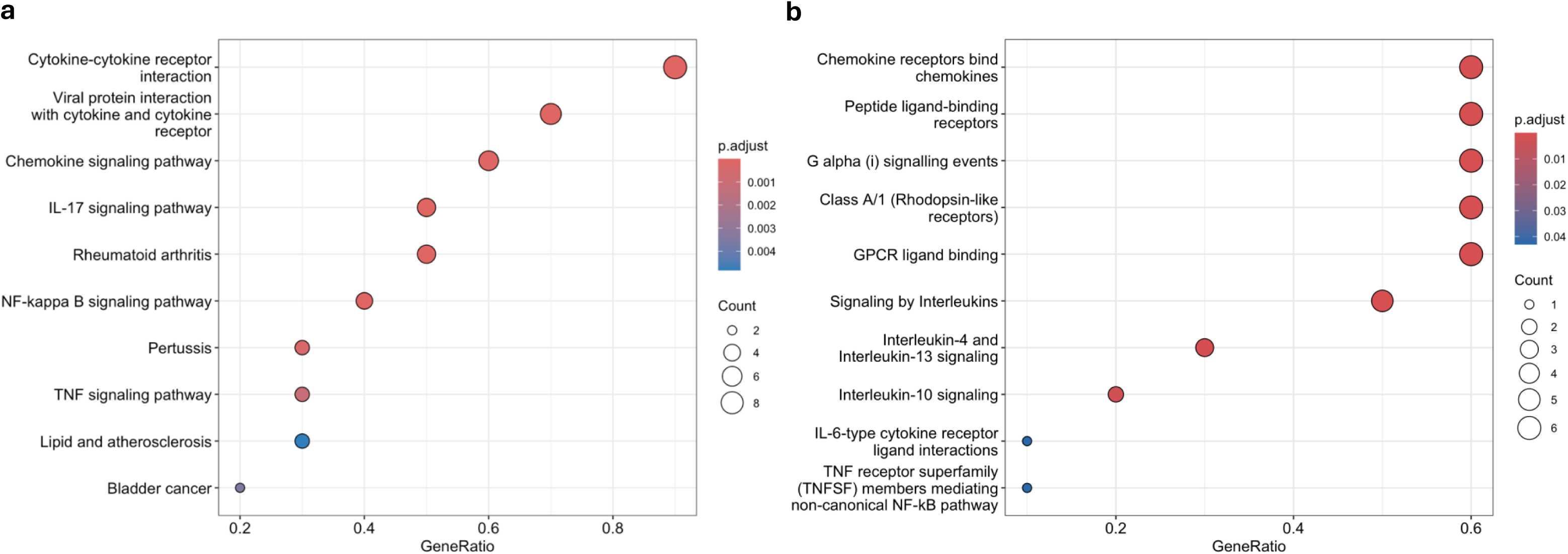
KEGG and Reactome pathway enrichment analyses. (a) KEGG pathway enrichment results. The x-axis represents the GeneRatio (the proportion of genes within each pathway that overlap with the list of differentially expressed proteins). Bubble size indicates the number of overlapping genes, while color corresponds to the adjusted p-value (FDR). (b) Reactome pathway enrichment results. The plot displays the enriched biological pathways identified by Reactome analysis. Bubble size reflects the number of genes associated with each path, and color denotes the adjusted p-value. Pathways with similar biological functions are clustered together.

Reactome pathway enrichment analysis was conducted to identify biological processes associated with the differentially expressed proteins between clusters. The analysis revealed significant enrichment of pathways primarily related to chemokine and cytokine receptor signaling and G protein–coupled receptor (GPCR)–mediated signaling (**Figure 4b**). The most significantly enriched pathway was “Chemokine receptors bind chemokines” (FDR = 9.11 × 10⁻¹¹; q = 3.55 × 10⁻¹¹), involving six of the ten proteins identified. Similarly, “Peptide ligand-binding receptors” (FDR = 8.22 × 10⁻⁸; q = 3.21 × 10⁻⁸), “G alpha (i) signaling events” (FDR = 8.52 × 10⁻⁷; q = 3.32 × 10⁻⁷), and “Class A/1 (Rhodopsin-like) receptors” (FDR = 8.56 × 10⁻⁷; q = 3.34 × 10⁻⁷) were among the top enriched pathways, underscoring a strong contribution of GPCR signaling. Downstream cytokine signaling cascades were also significantly enriched, including “Signaling by interleukins” (FDR = 1.09 × 10⁻⁴; q = 4.23 × 10⁻⁵), “Interleukin-4 and Interleukin-13 signaling” (FDR = 3.83 × 10⁻⁴; q = 1.49 × 10⁻⁴), and “Interleukin-10 signaling” (FDR = 2.56 × 10⁻³; q = 9.96 × 10⁻⁴). Additional pathways related to TNF receptor signaling, IL-6–type cytokine receptor interactions, and ATF4-mediated stress response were also identified at nominal significance levels. Complete Reactome enrichment results are provided in **Supplementary Material 1**.

## 4. DISCUSSION

Our findings demonstrate that patients with moderate-to-severe depression undergoing iTBS can be stratified into subgroups according to baseline inflammatory proteomic profiles, which are related with differences in treatment response. This extends prior evidence of biological heterogeneity in MDD observed in proteomic and multimodal studies (Milaneschi et al., 2020; Tang et al., 2025) and immune profiling (Hagenberg et al., 2025) by linking inflammation directly to neuromodulation outcomes. Two molecular phenotypes were identified, differing significantly in iTBS response but not in demographic or clinical confounders, suggesting that the variability may be related to immunological factors. Cluster 2 exhibited the highest response rate, whereas Cluster 1 contained the most non-responders, suggesting that baseline immune tone may be associated with therapeutic efficacy. Notably, the two clusters did not differ in baseline symptom severity or in mean change in total MADRS score. The principal clinical difference therefore lay in the likelihood of achieving a categorical treatment response rather than in average MADRS symptom reduction. When examining individual MADRS items descriptively, Cluster 2 also showed a tendency toward broader symptom improvement across several domains.

These findings support the view that MDD encompasses biologically (and to some extent, immunologically) distinct subtypes, one characterized by immune activation that may interfere with treatment response. While most depressed patients exhibit cytokine levels within the normal range, a substantial subset (estimated at one third) shows a pro-inflammatory phenotype (Miller and Raison, 2016; Osimo et al., 2019; Schmidt et al., 2016) linked to poorer antidepressant outcomes (Gadad et al., 2017; O’Brien et al., 2007; Strawbridge et al., 2015; Uher et al., 2014). The present findings indicate that such immune heterogeneity may similarly modulate response to iTBS, extending the relevance of inflammation beyond pharmacological treatment resistance.

Inflammatory cytokines appear to influence not only the onset and progression of depression but also responsiveness to treatment. Patients who respond to antidepressants typically exhibit normalization of cytokine levels, whereas non-responders maintain or increase inflammatory activity during therapy (Syed et al., 2018; Yilmaz et al., 2022). Elevated baseline IL-6, IL-8, and TNF-α have been consistently associated with reduced antidepressant efficacy (Gadad et al., 2017; Liu et al., 2020; Uher et al., 2014). Mechanistically, inflammation can also directly influence neural processes relevant to the antidepressant effects of iTBS. Pro-inflammatory cytokines such as IL-6, IL-8, and TNF-α, frequently elevated in patients with major depressive disorder (Krsek et al., 2024), are known to modulate synaptic plasticity (Lisowska, 2025; Singhal and Baune, 2017; Yirmiya and Goshen, 2011), processes critical for neuromodulation efficacy. Furthermore, TNF-α, IL-8, IFN-γ, and activation of the kynurenine pathway have been associated with alterations in white-matter microstructure in mood disorders (Poletti et al., 2024), suggesting that inflammatory dysregulation may extend beyond local synaptic effects to impact large-scale neural network integrity. Matrix metalloproteinases (e.g. MMP-1, which was elevated in the least responsive cluster) play dual roles in the central nervous system, mediating extracellular matrix remodeling essential for synaptic plasticity while, when dysregulated, contributing to blood–brain barrier breakdown and neuroinflammatory propagation (Hassamal, 2023; Huntley, 2012). Dysregulation of MMP signaling can therefore alter the neuronal microenvironment and limit synaptic remodeling, potentially reducing responsiveness to plasticity-inducing interventions (Alaiyed and Conant, 2019; Beroun et al., 2019).

Although our study assessed peripheral inflammatory proteins rather than direct central markers, peripheral immune activation can influence neuroinflammatory processes through several pathways, including cytokine signaling across the blood–brain barrier, endothelial and glial activation, and downstream effects on microglial function and kynurenine metabolism. In depression, such neuroimmune signaling has been linked to altered synaptic plasticity, impaired neurotrophic support, and disrupted network-level brain function, all of which are relevant to the proposed mechanism of iTBS. Preclinical studies further support this relationship by showing that depression-like behavior can be ameliorated through interventions that reduce neuroinflammation and related oxidative or nitrosative stress (Qiu et al., 2020; Ye et al., 2024).

From this perspective, the inflammatory profiles identified here may reflect not only systemic immune tone but also peripheral correlates of neuroinflammatory states that could reduce responsiveness to neuromodulation. Consistent with this interpretation, translational evidence indicates that heightened systemic inflammation is associated with diminished therapeutic responsiveness (Haroon et al., 2018), and reviews of rTMS in depression suggest that immune-inflammatory markers may be linked to neuromodulatory outcome (Rajkumar, 2023). Conversely, neuromodulation itself appears to exert immunoregulatory effects, reducing circulating pro-inflammatory cytokines and normalizing plasma MMP concentrations (Cirillo et al., 2023; Wang et al., 2022). In summary, these findings support the idea that iTBS may engage neuroimmune mechanisms and that specific inflammatory states could interfere with cortical plasticity and thereby attenuate treatment response.

The specific proteins that distinguish the poorer-outcome Cluster 1 provide further insight into the potential mechanisms underlying differential response to iTBS. The ten proteins that distinguish the two inflammatory clusters (MCP-4, CXCL6, IL-7, CXCL1, OSM, CXCL5, IL-8, TNFSF14, CXCL11, and MMP-1) are associated with enhanced chemotaxis, cytokine signaling, and extracellular matrix remodeling. These proteins are involved in immune pathways governing leukocyte recruitment and cytokine–cytokine receptor interactions, suggesting that non-responders may have a more pro-inflammatory profile. Consistent with this, enrichment analyses identified three major pathways with the strongest statistical support: cytokine–cytokine receptor interaction, chemokine receptor binding chemokines, and G-protein–coupled receptor (GPCR) signaling events. Together, these networks regulate immune cell trafficking, receptor-mediated signaling, and cellular responses to inflammatory stress. IL-8, a potent chemoattractant that drives neutrophil migration (Yoshimura et al., 1987), and related CXC chemokines (CXCL1, CXCL5, CXCL6, CXCL11) orchestrate leukocyte recruitment in the central nervous system (Milenkovic et al., 2019; Stuart et al., 2015) and have been linked to increased neuroinflammation and reduced antidepressant efficacy (Gasparini et al., 2022; Liu et al., 2020). MCP-4, a member of the MCP1–5 family, enhances macrophage chemotaxis and promotes chronic immune activation (Stuart et al., 2015). Oncostatin M (OSM), a pleiotropic IL-6–family cytokine, promotes endothelial activation and leukocyte transmigration (Han et al., 2023; Modur et al., 1997) and has been implicated in pro-inflammatory and mood-related signaling cascades (Franzen et al., 2020; Knight, 2022; Richards, 2013; Tanaka and Miyajima, 2003). IL-7, which regulates T-cell proliferation and survival (Chen et al., 2021), may further modulate neuroimmune crosstalk, while MMP-1, a matrix metalloproteinase central to extracellular matrix remodeling (Pardo and Selman, 2005), can affect synaptic integrity and blood–brain-barrier permeability when dysregulated (Hassamal, 2023; Huntley, 2012). Collectively, these findings suggest that upregulated cytokine and chemokine signaling, coupled with aberrant matrix remodeling, may be associated with lower iTBS efficacy, whereas lower inflammatory tone may be associated with better response.

The principal strength of this study lies in its data-driven identification of inflammatory subgroups in moderate-to-severe depression, linking baseline immune profiles to differential iTBS outcomes and enrichment of cytokine and interleukin signaling pathways. However, several limitations must be considered. The exploratory design precludes causal inference, and findings should be regarded as hypothesis-generating pending replication in larger, longitudinal cohorts. In particular, the relatively small number of participants within each cluster (Cluster 1: n = 28; Cluster 2: n = 26) limits statistical precision and calls for cautious interpretation of subgroup estimates, even though the clustering solution itself showed high bootstrap stability. Analyses were restricted to baseline protein expression; inclusion of post-treatment data would clarify how inflammatory signatures evolve with iTBS. Additionally, the lack of comparator arms (e.g., pharmacotherapy or ECT) constrains conclusions about the specificity of these immune patterns to neuromodulation. While the Olink® Inflammation panel focuses on inflammation-related proteins, which aligns well with our a priori interest based on previous findings, future work using broader proteomic approaches may help to uncover additional biological pathways associated with iTBS outcomes and treatment response. Furthermore, the modest number of significant proteins and the presence of potential confounders (such as lifestyle and psychosocial factors) further limit interpretability. Although participants differed in concomitant medication, sensitivity analyses suggested that neither medication category nor medication load explained the observed association between inflammatory cluster membership and treatment response. Finally, since all participants presented with moderate-to-severe depression, the generalizability of these findings to milder forms of depression remains uncertain. Nevertheless, using a conservative |ΔNPX| ≥ 1 threshold (≈2-fold change on the log₂ scale) ensured robust, interpretable results, and all significant proteins achieved power > 0.99 (**see Supplementary Material 1**).

In conclusion, baseline inflammatory proteomic profiles are associated with subgroups of patients with moderate-to-severe depression who differ in their clinical response to iTBS. Cluster 1, defined by elevated cytokine- and chemokine-related markers, showed poorer outcomes, whereas Cluster 2, characterized by lower baseline immune activity, exhibited better improvement. Cytokine signaling pathways regulate proliferation, apoptosis, and immune activation (Voßhenrich and Santo, 2002), and excessive activation has been associated with reduced cortical adaptability. These findings support the concept that immune dysregulation contributes to treatment resistance and may serve as a biological modifier of neuromodulatory efficacy. Future research should validate these results in larger cohorts, incorporate post-treatment proteomics and neuroimaging, and test whether the ten significant markers predict responsiveness across treatment modalities. Ultimately, such research could guide precision-based interventions, including anti-inflammatory augmentation strategies, for patients with depression who exhibit an inflammatory profile.

## Data Availability

The data supporting the findings of this study are not publicly available due to European Union data protection regulations. However, data may be made available upon reasonable request from the corresponding author, provided appropriate legal and ethical conditions are met.

## 5. ACKNOWLEGEMENTS

This work was supported by the German Federal Ministry of Education and Research (*Bundesministerium für Bildung und Forschung, BMBF: 01 ZX 1507*, ‘‘PreNeSt - e:Med’’) and the to the foundation *Immunität und Seele (*https://immunitaet-und-seele.de/). The authors thank the team of Dr. Nicole Ziegler (Fraunhofer Institute for Translational Medicine and Pharmacology, Frankfurt am Main, Germany) for their assistance in analyzing blood samples using proteomics panels.

## 6. CONFLICTS OF INTEREST AND OTHER STATEMENTS

**Ethics Approval and Consent to Participate**: The study protocol was conducted in accordance with the Declaration of Helsinki and was approved by the Ethics Committee of the University Medical Center Göttingen (UMG). All participants provided both verbal and written informed consent after receiving a full explanation of the study procedures.

**Consent for Publication**: Not applicable.

**Availability of Data and Materials**: The data supporting the findings of this study are not publicly available due to European Union data protection regulations. However, data may be made available upon reasonable request from the corresponding author, provided appropriate legal and ethical conditions are met.

**Statement**: During the preparation of this manuscript, the authors used Grammarly Inc., Nature Research Assistant, and ChatGPT to assist with language refinement and grammar correction. Elicit was used to support the literature search. All content was critically reviewed and edited by the authors, who take full responsibility for the final version of the manuscript.

**Competing Interests**: The authors declare that they have no competing interests.

**Funding**: This project received financial support from the German Federal Ministry of Education and Research (*Bundesministerium für Bildung und Forschung, BMBF: 01 ZX 1507*, ‘‘PreNeSt - e:Med’’) and the foundation *Immunität und Seele* (Prof. Dr. Norbert Müller https://immunitaet-und-seele.de/). The funders had no role in the study design, data collection, analysis, interpretation, or manuscript preparation.

**Employment**: None of the authors was employed or contracted by any organization that could gain or lose financially from the publication of this manuscript.

**Personal Financial Interests**: The authors did not receive any form of financial compensation (e.g., salaries, stocks, consulting fees) from commercial entities related to this research.

**Declarations of Interest**: Bruno Pedraz acknowledges receiving a Seed Money for Research grant from the Central Institute of Mental Health and receiving a grant from the foundation *Immunität und Seele*. Roberto Goya-Maldonado acknowledges receiving a grant from the German Federal Ministry of Education and Research (*Bundesministerium für Bildung und Forschung, BMBF: 01 ZX 1507*, ‘‘PreNeSt-e:Med’’).

## 1 SUPPLEMENTAL MATERIAL

**Supplementary Figure 1.**
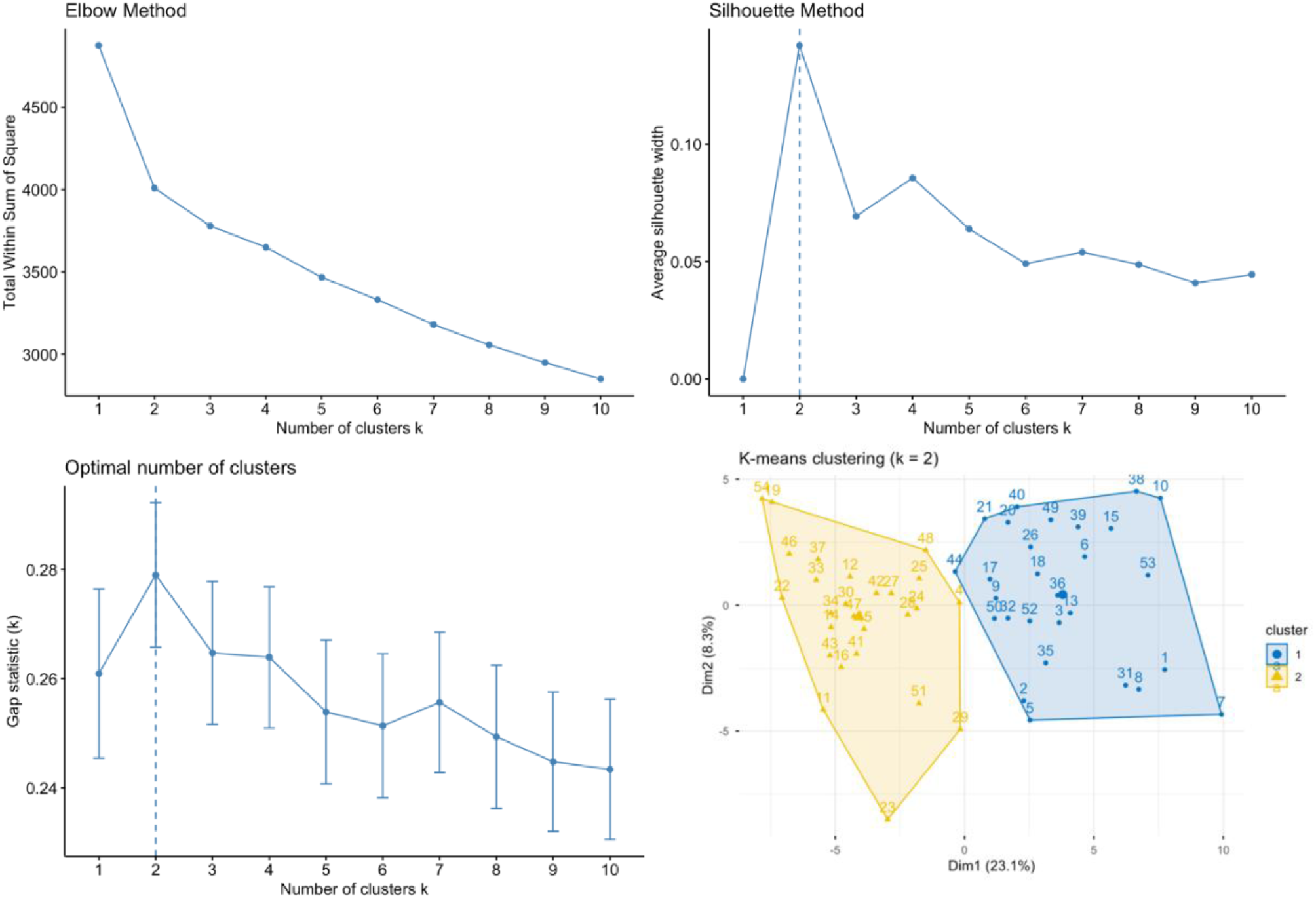
Exploratory cluster validity diagnostics using the Elbow, Gap Statistics, and Silhouette methods. All metrics were computed for k values ranging from 1 to 10, based on the standardized expression of inflammatory proteins. All methods suggest a cluster solution of k = 2.

**Table S1.**
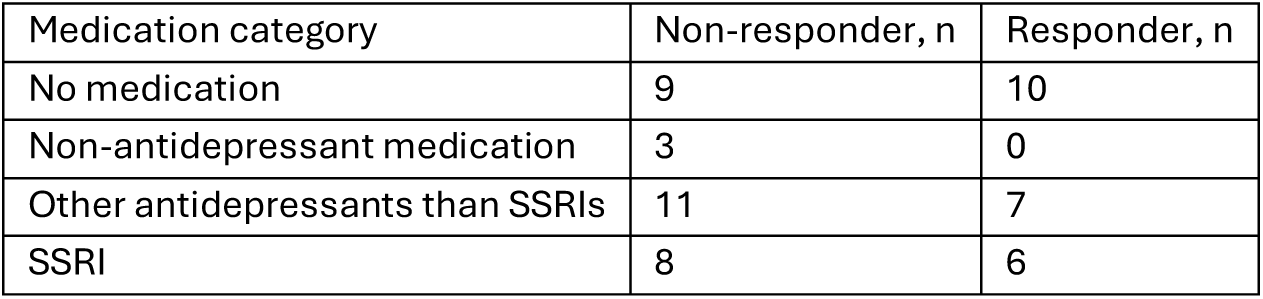
Distribution of medication category across inflammatory clusters and responder groups.

**Table S2.**
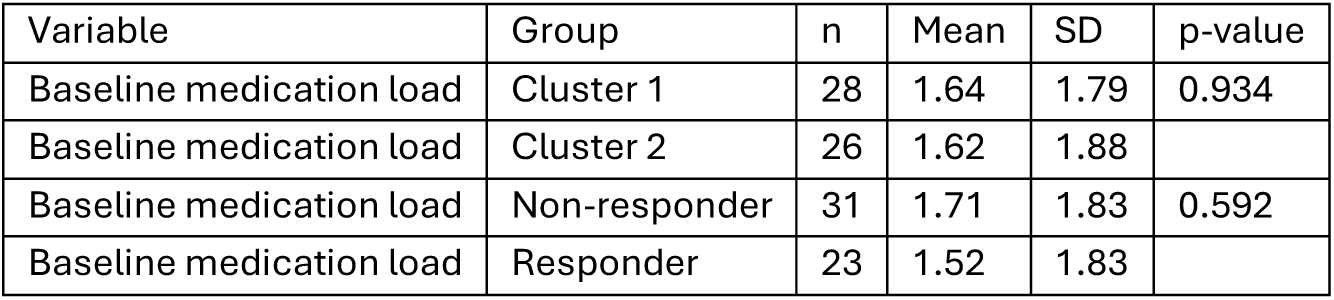
Baseline Medication load by cluster and responder status. Medication-load comparisons were performed using Wilcoxon rank-sum tests. P-values are from Wilcoxon rank-sum tests.

**Table S3.**
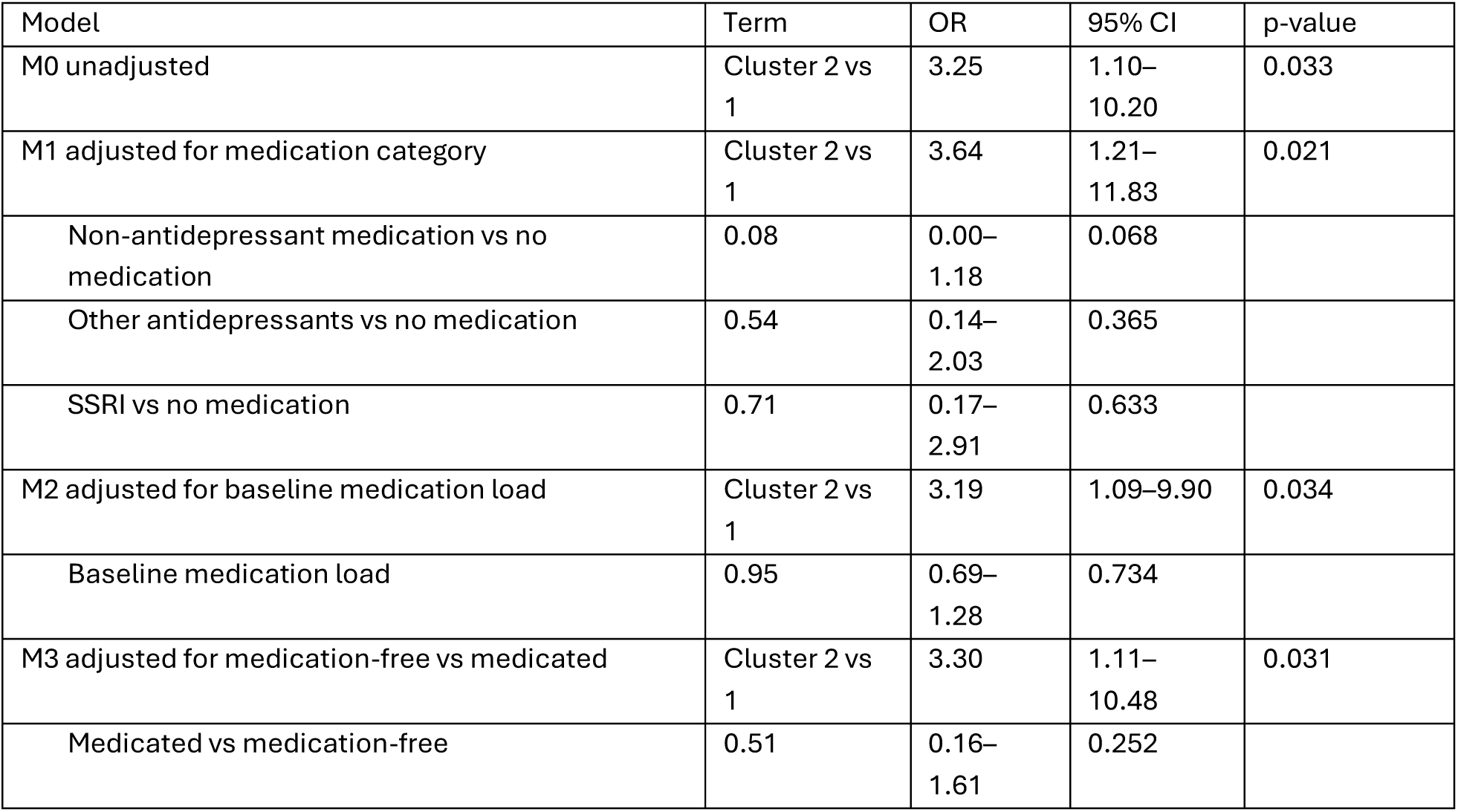
Firth penalized logistic regression models examining medication-related confounding of the association between inflammatory cluster membership and treatment response. Odds ratios (ORs), 95% confidence intervals (CIs), and p-values are from Firth penalized logistic regression models with responder status as the dependent variable. Medication was coded as no medication (at all), non-antidepressant medication, antidepressants other than SSRIs, and SSRIs.

**Table S4.**
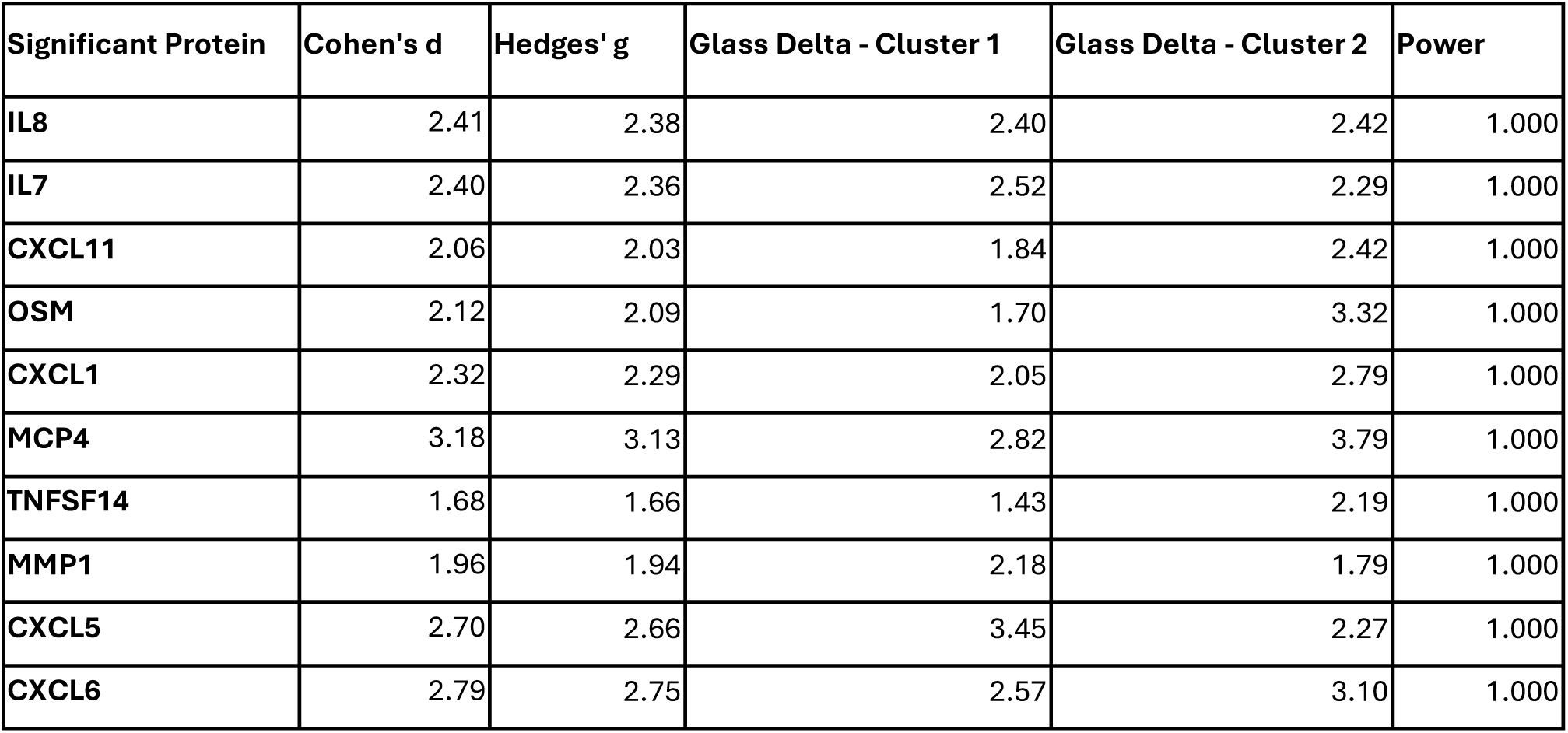
Proteins showing significant differences in expression between Cluster 1 and Cluster 2 (|ΔNPX| ≥ 1.0 and FDR < 0.05). Effect sizes are reported as Cohen’s d, Hedges’ g, and Glass’s Δ for each cluster. All proteins achieved post-hoc power > 0.999.

**Table S5.**
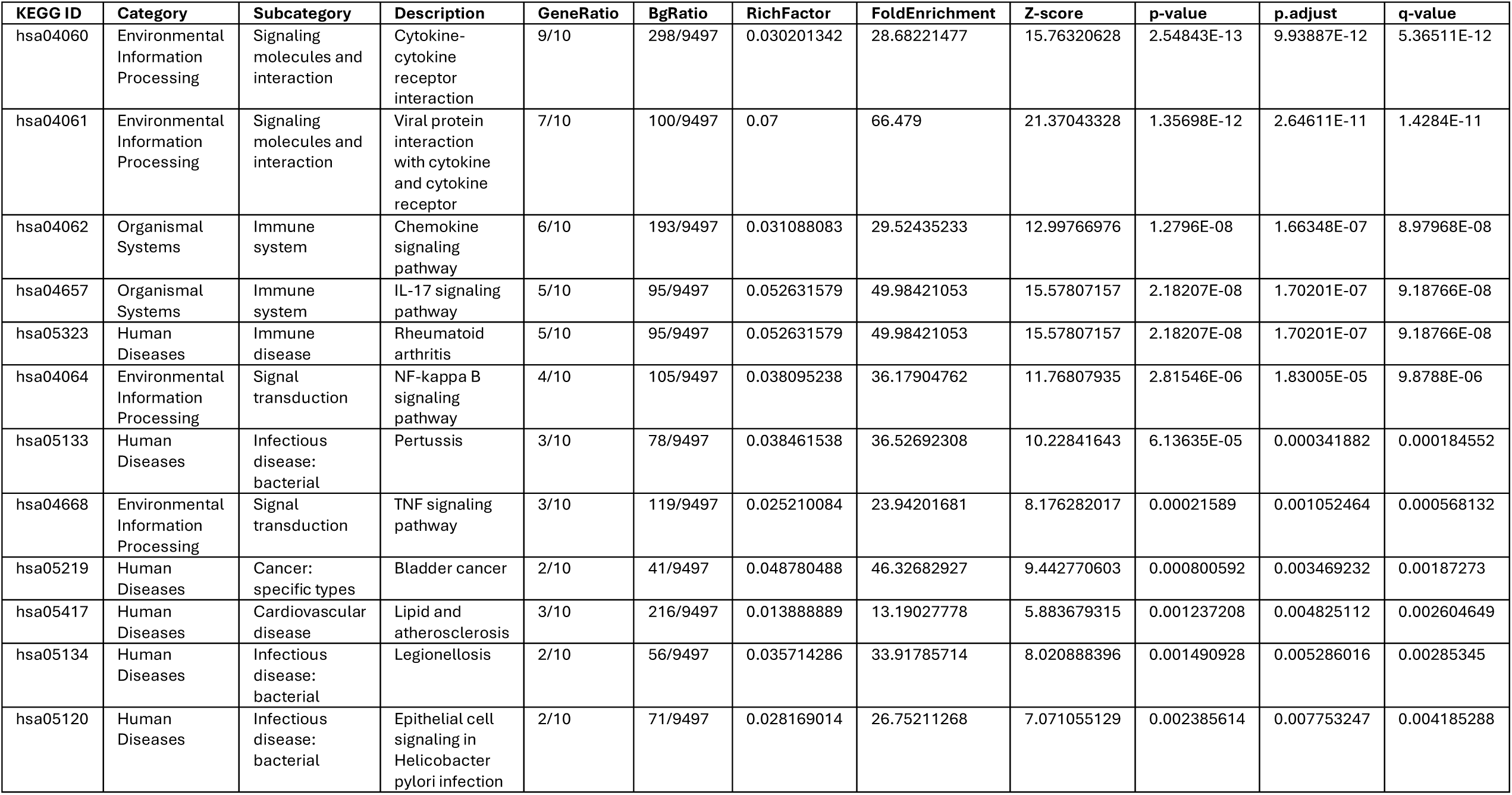
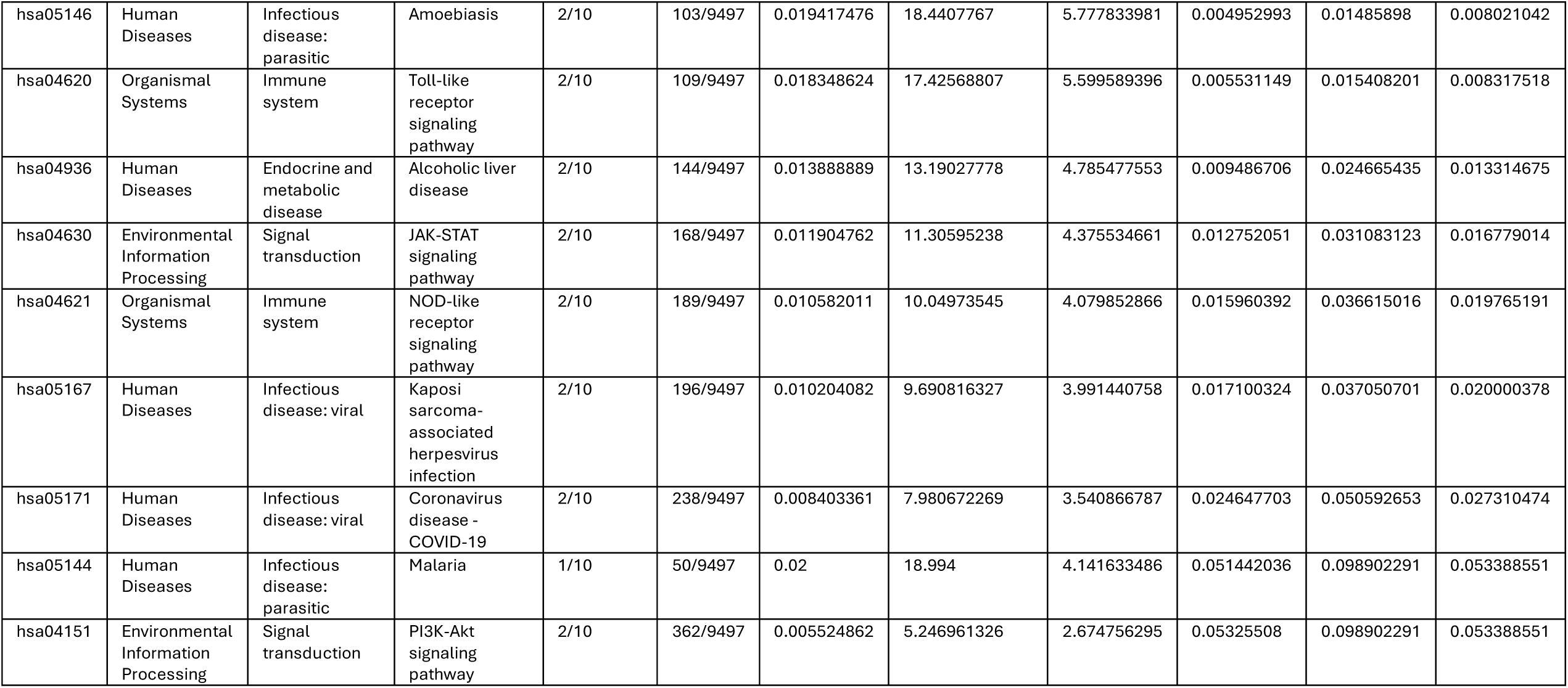
KEGG Pathway Enrichment Analysis - This table presents the results of KEGG pathway enrichment analysis performed on the differentially expressed proteins. Pathways are identified by KEGG ID and grouped by category. The columns display the gene ratio, background ratio, enrichment scores, p-values, and the corresponding genes.

**Table S6.**
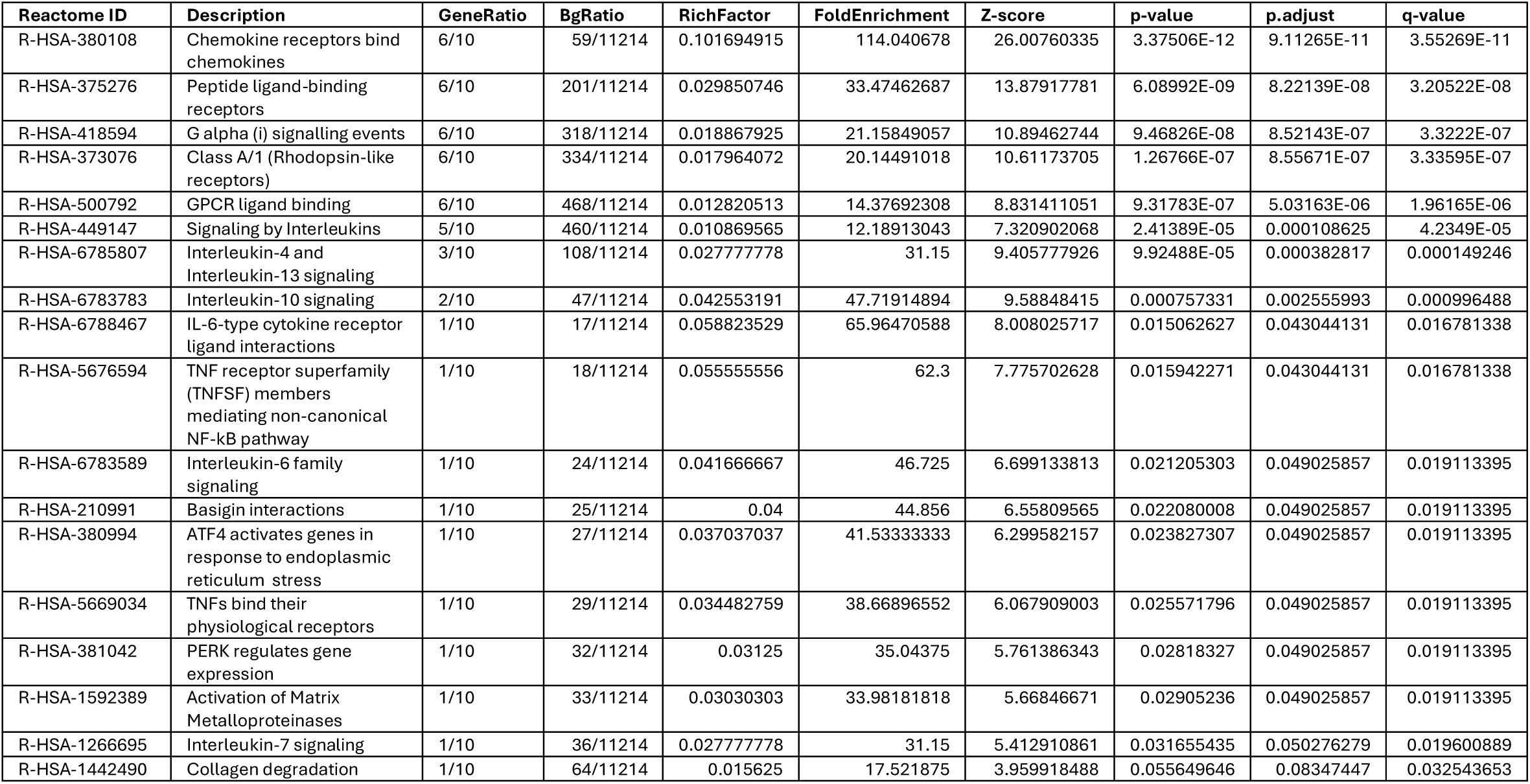
Reactome Pathway Enrichment Analysis - This table presents the results of Reactome pathway enrichment analysis. The analysis identifies biological pathways significantly enriched among differentially expressed proteins. Key metrics include gene ratios, enrichment factors, and statistical significance values.

## 2: SUPPLEMENTAL MATERIAL

**Table S1.**
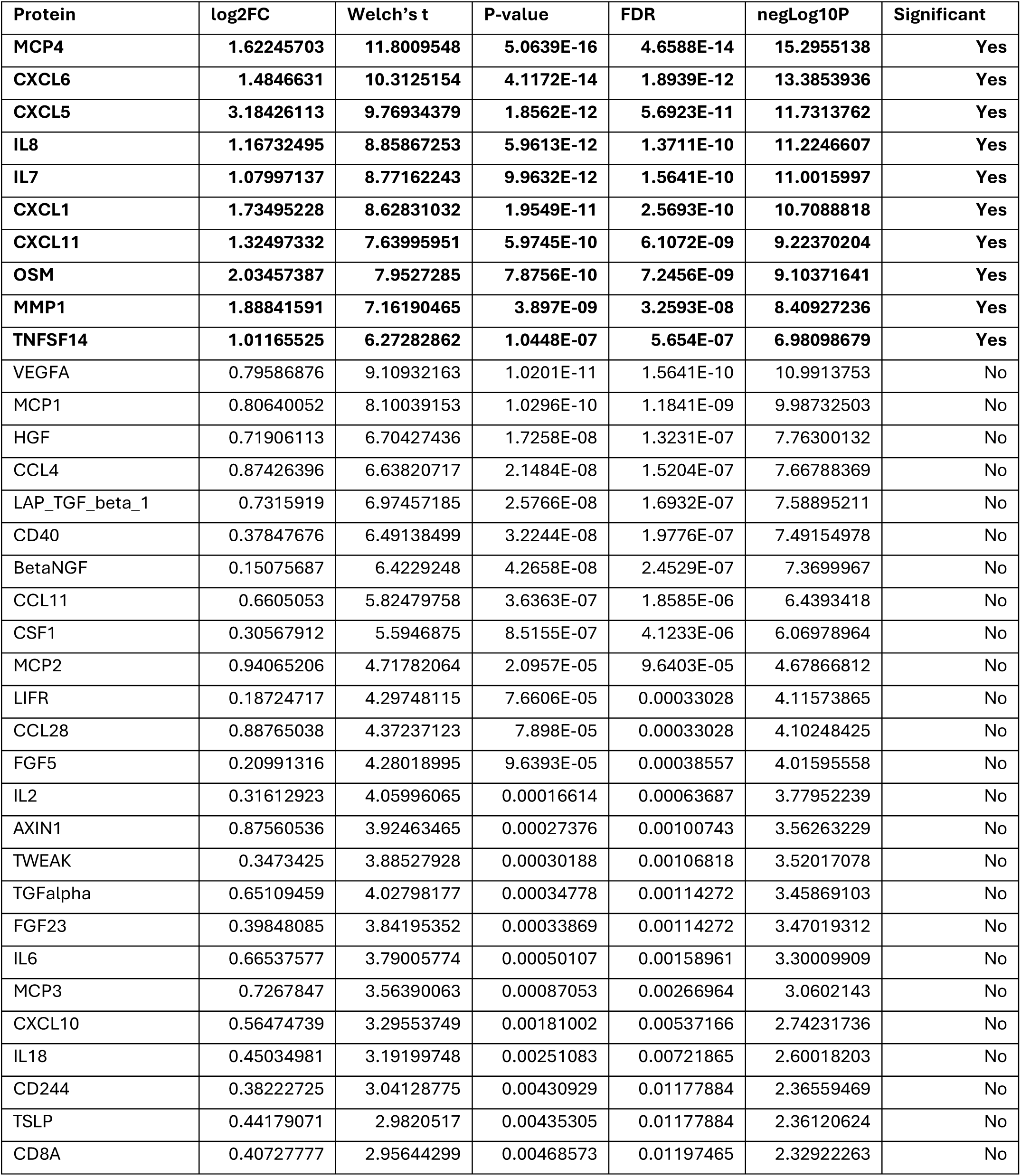
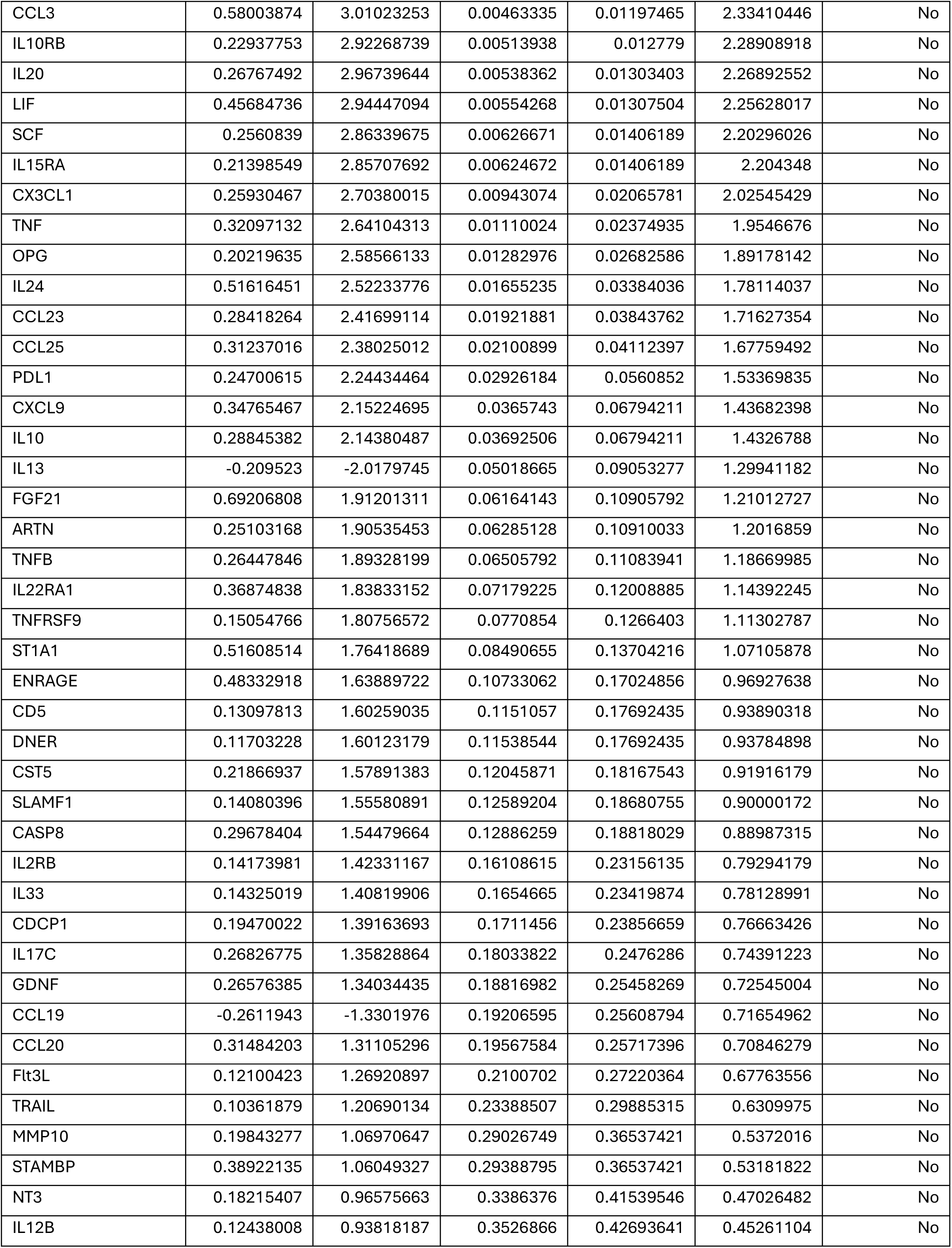
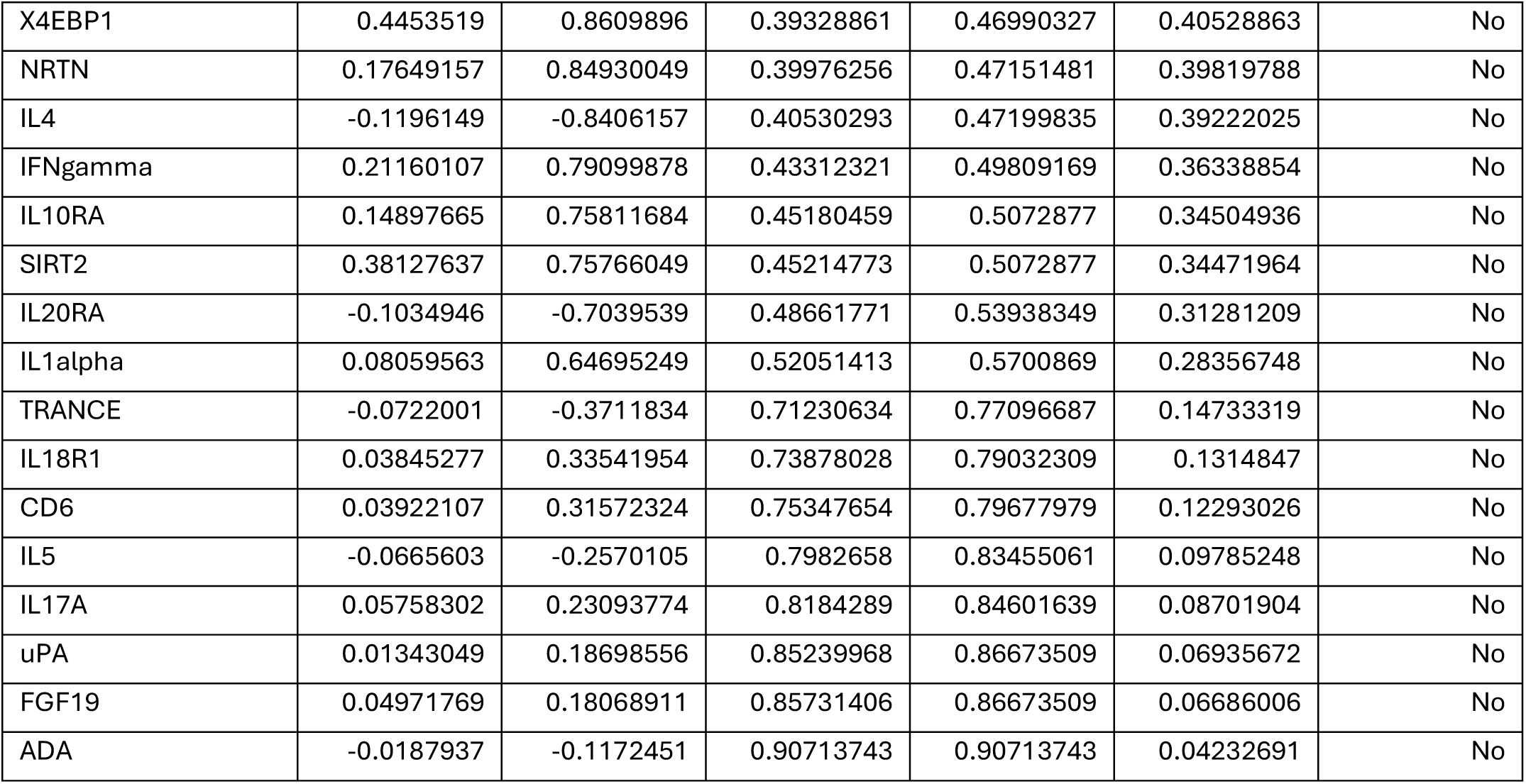
Differential inflammatory protein expression between Clusters 1 and 2. Log₂ fold change (log2FC), t-statistics, p-values, false discovery rate (FDR, Benjamini–Hochberg correction), and −log₁₀(p) values are shown. Proteins meeting both thresholds (FDR < 0.05 and |log₂FC| ≥ 1) are marked as significant.

## ABBREVIATION LIST

4E-BP1: Eukaryotic Translation Initiation Factor 4E-Binding Protein 1
ADA: Adenosine Deaminase
ARTN: Artemin
AXIN1: Axis Inhibition Protein 1
Beta-NGF: Beta Nerve Growth Factor
BMI: Body Mass Index
CASP-8: Caspase 8
CCL11: C-C Motif Chemokine Ligand 11 (Eotaxin-1)
CCL19: C-C Motif Chemokine Ligand 19
CCL20: C-C Motif Chemokine Ligand 20
CCL23: C-C Motif Chemokine Ligand 23
CCL25: C-C Motif Chemokine Ligand 25
CCL28: C-C Motif Chemokine Ligand 28
CCL3: C-C Motif Chemokine Ligand 3
CCL4: C-C Motif Chemokine Ligand 4
CD244: CD244 Molecule (2B4)
CD40: CD40 Molecule
CD5: CD5 Molecule
CD6: CD6 Molecule
CD8A: CD8a Molecule
CDCP1: CUB Domain Containing Protein 1
CNS: Central Nervous System
CRP: C-Reactive Protein
CSF-1: Colony Stimulating Factor 1
CST5: Cystatin D
CX3CL1: CX3C Motif Chemokine Ligand 1 (Fractalkine)
CXC: C-X-C Motif Chemokine Family
CXCL: C-X-C Motif Chemokine Ligand (e.g., CXCL1, CXCL5, CXCL6, CXCL11)
CXCL1: C-X-C Motif Chemokine Ligand 1
CXCL10: C-X-C Motif Chemokine Ligand 10
CXCL11: C-X-C Motif Chemokine Ligand 11
CXCL5: C-X-C Motif Chemokine Ligand 5
CXCL6: C-X-C Motif Chemokine Ligand 6
CXCL9: C-X-C Motif Chemokine Ligand 9
DEPs: Differentially Expressed Proteins
DLPFC: Dorsolateral Prefrontal Cortex
DNER: Delta/Notch-Like EGF Repeat Containing
DSM-5: Diagnostic and Statistical Manual of Mental Disorders, Fifth Edition
ECT: Electroconvulsive Therapy
EDTA: Ethylenediaminetetraacetic Acid
EN-RAGE: Extracellular Newly Identified RAGE-Binding Protein (S100A12)
FDA U.S.: Food and Drug Administration
FDR: False Discovery Rate
FGF-19: Fibroblast Growth Factor 19
FGF-21: Fibroblast Growth Factor 21
FGF-23: Fibroblast Growth Factor 23
FGF-5: Fibroblast Growth Factor 5
Flt3L: FMS-like Tyrosine Kinase 3 Ligand
GDNF: Glial Cell Line-Derived Neurotrophic Factor
gp130: Glycoprotein 130
GPCR: G-Protein–Coupled Receptor
HGF: Hepatocyte Growth Factor
HPA: Hypothalamic–Pituitary–Adrenal Axis
IFNγ: Interferon Gamma
IL: Interleukin (e.g., IL-6, IL-8, IL-10, IL-13)
IL10RA: Interleukin 10 Receptor Subunit Alpha
IL10RB: Interleukin 10 Receptor Subunit Beta
IL12B: Interleukin 12 Subunit Beta
IL15RA: Interleukin 15 Receptor Alpha
IL17A: Interleukin 17A
IL17C: Interleukin 17C
IL18R1: Interleukin 18 Receptor 1
IL1α: Interleukin 1 Alpha
IL1β: Interleukin 1 Beta
IL20: Interleukin 20
IL20RA: Interleukin 20 Receptor Alpha
IL22RA1: Interleukin 22 Receptor Subunit Alpha 1
IL24: Interleukin 24
IL2RB: Interleukin 2 Receptor Subunit Beta
IL6: Interleukin 6
IL10: Interleukin 10
IL13: Interleukin 13
IL18: Interleukin 18
IL2: Interleukin 2
IL33: Interleukin 33
IL4: Interleukin 4
IL5: Interleukin 5
IL7: Interleukin 7
IL8: Interleukin 8
ITAC: Interferon-inducible T-cell Alpha Chemoattractant
iTBS: Intermittent Theta Burst Stimulation
JAK/STAT: Janus Kinase/Signal Transducer and Activator of Transcription Pathway
KEGG: Kyoto Encyclopedia of Genes and Genomes
LAP: TGF-beta-1 Latency-Associated Peptide of TGF-β1
LIF: Leukemia Inhibitory Factor
LIF-R: Leukemia Inhibitory Factor Receptor
MADRS: Montgomery–Åsberg Depression Rating Scale
MAPK: Mitogen-Activated Protein Kinase
MCP: Monocyte Chemotactic Protein
MCP-1: Monocyte Chemoattractant Protein 1 (CCL2)
MCP-2: Monocyte Chemoattractant Protein 2 (CCL8)
MCP-3: Monocyte Chemoattractant Protein 3 (CCL7)
MCP-4: Monocyte Chemoattractant Protein 4 (CCL13)
MDD: Major Depressive Disorder
MMP: Matrix Metalloproteinase (e.g., MMP-1)
MMP-1: Matrix Metallopeptidase 1
MMP-10: Matrix Metallopeptidase 10
MRI: Magnetic Resonance Imaging
NF-κB: Nuclear Factor Kappa-Light-Chain-Enhancer of Activated B Cells
NPX: Normalized Protein Expression (Olink Platform Metric)
NRTN: Neurturin
NT-3: Neurotrophin-3
OPG: Osteoprotegerin (TNFRSF11B)
OSM: Oncostatin M
PD-L1: Programmed Death-Ligand 1 (CD274)
PEA: Proximity Extension Assay
pMADRS: Percentage Change in MADRS Item
RMT: Resting Motor Threshold
rTMS: Repetitive Transcranial Magnetic Stimulation
SCF: Stem Cell Factor (KITLG)
SCID-5-CV: Structured Clinical Interview for DSM-5 Disorders – Clinical Version
SD: Standard Deviation
SIRT2: Sirtuin 2
SLAMF1: SLAM Family Member 1
ST1A1: Sulfotransferase Family 1A Member 1
STAMBP: STAM Binding Protein
STAT3: Signal Transducer and Activator of Transcription 3
tENS: Transcutaneous Electrical Nerve Stimulation
TGF-alpha: Transforming Growth Factor Alpha
TNF: Tumor Necrosis Factor
TNF-α: Tumor Necrosis Factor Alpha
TNFB: Tumor Necrosis Factor Beta (Lymphotoxin Alpha)
TNFRSF9: TNF Receptor Superfamily Member 9 (4-1BB/CD137)
TNFSF14: TNF Superfamily Member 14 (LIGHT)
TRAIL: TNF-Related Apoptosis-Inducing Ligand (TNFSF10)
TRANCE: TNF-Related Activation-Induced Cytokine (RANKL/TNFSF11)
TSLP: Thymic Stromal Lymphopoietin
TWEAK: TNF-Like Weak Inducer of Apoptosis (TNFSF12)
uPA: Urokinase-type Plasminogen Activator
VEGFA: Vascular Endothelial Growth Factor A
ΔMADRS: Change in MADRS Score (post-treatment minus baseline)

