## Supplemental Material for "Baseline Inflammatory Profiles in Moderate-to-Severe Depression and Differential Response to Intermittent Theta-Burst Stimulation"

**SUPPLEMENTAL MATERIAL 1**

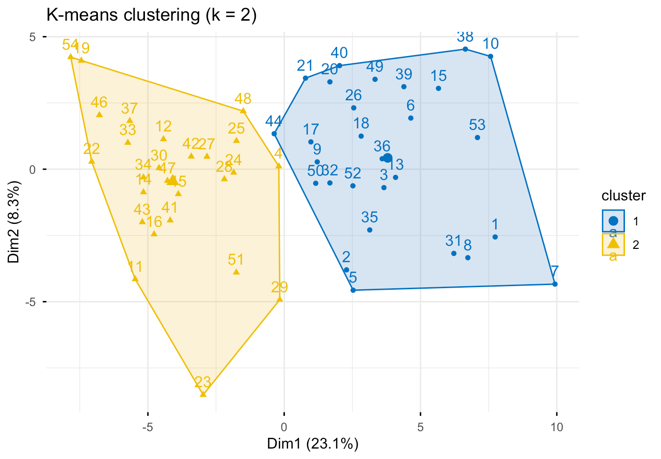

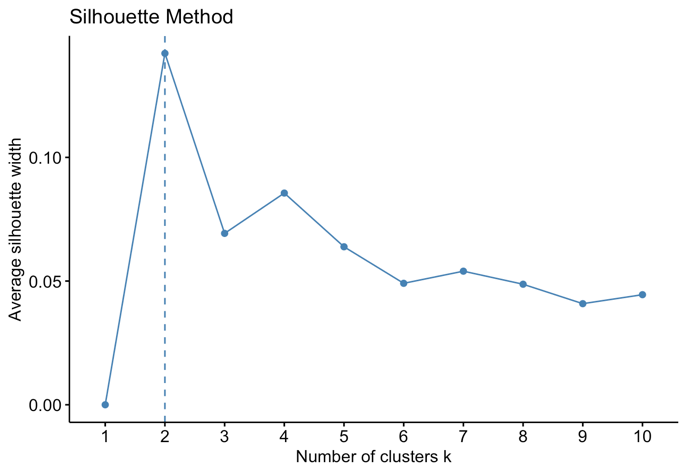

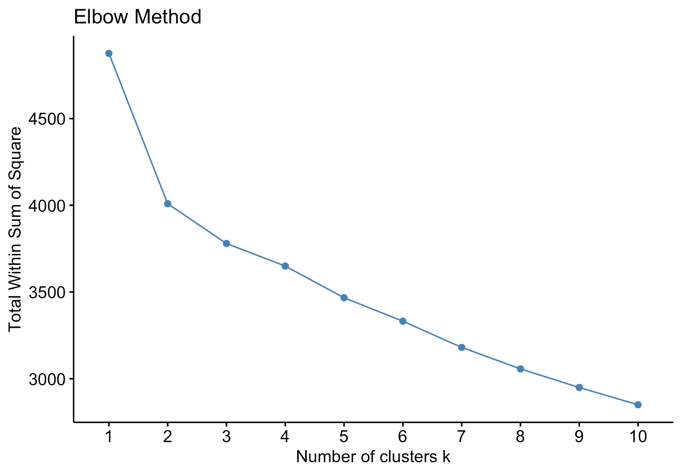

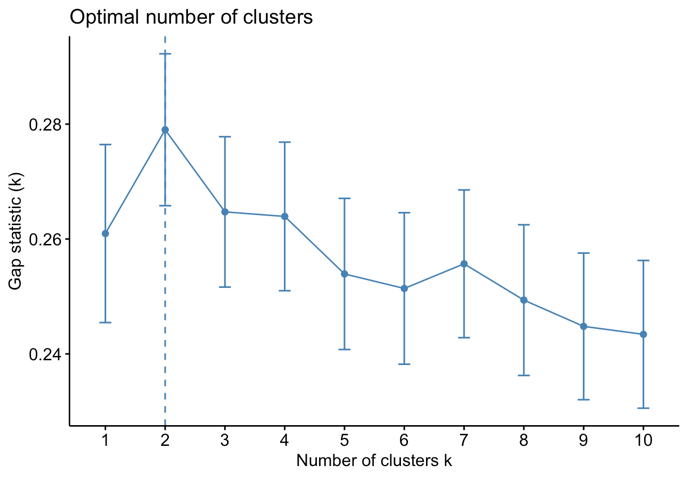

***Supplementary Figure 1.*** *Exploratory cluster validity diagnostics using the Elbow, Gap Statistics, and Silhouette methods. All metrics were computed for k values ranging from 1 to 10, based on the standardized expression of inflammatory proteins. All methods suggest a cluster solution of k = 2.*

***Table S1.*** *Distribution of medication category across inflammatory clusters and responder groups*

| Medication category | Non-responder, n | Responder, n |
| --- | --- | --- |
| No medication | 9 | 10 |
| Non-antidepressant medication | 3 | 0 |
| Other antidepressants than SSRIs | 11 | 7 |
| SSRI | 8 | 6 |

***Table S2.*** *Baseline Medication load by cluster and responder status. Medication-load comparisons were performed using Wilcoxon rank-sum tests. P-values are from Wilcoxon rank-sum tests.*

| Variable | Group | n | Mean | SD | p-value |
| --- | --- | --- | --- | --- | --- |
| Baseline medication load | Cluster 1 | 28 | 1.64 | 1.79 | 0.934 |
| Baseline medication load | Cluster 2 | 26 | 1.62 | 1.88 |  |
| Baseline medication load | Non-responder | 31 | 1.71 | 1.83 | 0.592 |
| Baseline medication load | Responder | 23 | 1.52 | 1.83 |  |

***Table S3.*** *Firth penalized logistic regression models examining medication-related confounding of the association between inflammatory cluster membership and treatment response. Odds ratios (ORs), 95% confidence intervals (CIs), and p-values are from Firth penalized logistic regression models with responder status as the dependent variable. Medication was coded as no medication (at all), non-antidepressant medication, antidepressants other than SSRIs, and SSRIs.*

| Model | Term | OR | 95% CI | p-value |
| --- | --- | --- | --- | --- |
| M0 unadjusted | Cluster 2 vs 1 | 3.25 | 1.10–10.20 | 0.033 |
| M1 adjusted for medication category | Cluster 2 vs 1 | 3.64 | 1.21–11.83 | 0.021 |
| Non-antidepressant medication vs no medication | 0.08 | 0.00–1.18 | 0.068 |  |
| Other antidepressants vs no medication | 0.54 | 0.14–2.03 | 0.365 |  |
| SSRI vs no medication | 0.71 | 0.17–2.91 | 0.633 |  |
| M2 adjusted for baseline medication load | Cluster 2 vs 1 | 3.19 | 1.09–9.90 | 0.034 |
| Baseline medication load | 0.95 | 0.69–1.28 | 0.734 |  |
| M3 adjusted for medication-free vs medicated | Cluster 2 vs 1 | 3.30 | 1.11–10.48 | 0.031 |
| Medicated vs medication-free | 0.51 | 0.16–1.61 | 0.252 |  |

***Table S4.*** *Proteins showing significant differences in expression between Cluster 1 and Cluster 2 (|ΔNPX| ≥ 1.0 and FDR < 0.05). Effect sizes are reported as Cohen’s d, Hedges’ g, and Glass’s Δ for each cluster. All proteins achieved post-hoc power > 0.999.*

| **Significant Protein** | **Cohen's d** | **Hedges' g** | **Glass Delta - Cluster 1** | **Glass Delta - Cluster 2** | **Power** |
| --- | --- | --- | --- | --- | --- |
| **IL8** | 2.41 | 2.38 | 2.40 | 2.42 | 1.000 |
| **IL7** | 2.40 | 2.36 | 2.52 | 2.29 | 1.000 |
| **CXCL11** | 2.06 | 2.03 | 1.84 | 2.42 | 1.000 |
| **OSM** | 2.12 | 2.09 | 1.70 | 3.32 | 1.000 |
| **CXCL1** | 2.32 | 2.29 | 2.05 | 2.79 | 1.000 |
| **MCP4** | 3.18 | 3.13 | 2.82 | 3.79 | 1.000 |
| **TNFSF14** | 1.68 | 1.66 | 1.43 | 2.19 | 1.000 |
| **MMP1** | 1.96 | 1.94 | 2.18 | 1.79 | 1.000 |
| **CXCL5** | 2.70 | 2.66 | 3.45 | 2.27 | 1.000 |
| **CXCL6** | 2.79 | 2.75 | 2.57 | 3.10 | 1.000 |

### ***Table S5.*** *KEGG Pathway Enrichment Analysis - This table presents the results of KEGG pathway enrichment analysis performed on the differentially expressed proteins. Pathways are identified by KEGG ID and grouped by category. The columns display the gene ratio, background ratio, enrichment scores, p-values, and the corresponding genes.*

| **KEGG ID** | **Category** | **Subcategory** | **Description** | **GeneRatio** | **BgRatio** | **RichFactor** | **FoldEnrichment** | **Z-score** | **p-value** | **p.adjust** | **q-value** |
| --- | --- | --- | --- | --- | --- | --- | --- | --- | --- | --- | --- |
| hsa04060 | Environmental Information Processing | Signaling molecules and interaction | Cytokine-cytokine receptor interaction | 9/10 | 298/9497 | 0.030201342 | 28.68221477 | 15.76320628 | 2.54843E-13 | 9.93887E-12 | 5.36511E-12 |
| hsa04061 | Environmental Information Processing | Signaling molecules and interaction | Viral protein interaction with cytokine and cytokine receptor | 7/10 | 100/9497 | 0.07 | 66.479 | 21.37043328 | 1.35698E-12 | 2.64611E-11 | 1.4284E-11 |
| hsa04062 | Organismal Systems | Immune system | Chemokine signaling pathway | 6/10 | 193/9497 | 0.031088083 | 29.52435233 | 12.99766976 | 1.2796E-08 | 1.66348E-07 | 8.97968E-08 |
| hsa04657 | Organismal Systems | Immune system | IL-17 signaling pathway | 5/10 | 95/9497 | 0.052631579 | 49.98421053 | 15.57807157 | 2.18207E-08 | 1.70201E-07 | 9.18766E-08 |
| hsa05323 | Human Diseases | Immune disease | Rheumatoid arthritis | 5/10 | 95/9497 | 0.052631579 | 49.98421053 | 15.57807157 | 2.18207E-08 | 1.70201E-07 | 9.18766E-08 |
| hsa04064 | Environmental Information Processing | Signal transduction | NF-kappa B signaling pathway | 4/10 | 105/9497 | 0.038095238 | 36.17904762 | 11.76807935 | 2.81546E-06 | 1.83005E-05 | 9.8788E-06 |
| hsa05133 | Human Diseases | Infectious disease: bacterial | Pertussis | 3/10 | 78/9497 | 0.038461538 | 36.52692308 | 10.22841643 | 6.13635E-05 | 0.000341882 | 0.000184552 |
| hsa04668 | Environmental Information Processing | Signal transduction | TNF signaling pathway | 3/10 | 119/9497 | 0.025210084 | 23.94201681 | 8.176282017 | 0.00021589 | 0.001052464 | 0.000568132 |
| hsa05219 | Human Diseases | Cancer: specific types | Bladder cancer | 2/10 | 41/9497 | 0.048780488 | 46.32682927 | 9.442770603 | 0.000800592 | 0.003469232 | 0.00187273 |
| hsa05417 | Human Diseases | Cardiovascular disease | Lipid and atherosclerosis | 3/10 | 216/9497 | 0.013888889 | 13.19027778 | 5.883679315 | 0.001237208 | 0.004825112 | 0.002604649 |
| hsa05134 | Human Diseases | Infectious disease: bacterial | Legionellosis | 2/10 | 56/9497 | 0.035714286 | 33.91785714 | 8.020888396 | 0.001490928 | 0.005286016 | 0.00285345 |
| hsa05120 | Human Diseases | Infectious disease: bacterial | Epithelial cell signaling in Helicobacter pylori infection | 2/10 | 71/9497 | 0.028169014 | 26.75211268 | 7.071055129 | 0.002385614 | 0.007753247 | 0.004185288 |
| hsa05146 | Human Diseases | Infectious disease: parasitic | Amoebiasis | 2/10 | 103/9497 | 0.019417476 | 18.4407767 | 5.777833981 | 0.004952993 | 0.01485898 | 0.008021042 |
| hsa04620 | Organismal Systems | Immune system | Toll-like receptor signaling pathway | 2/10 | 109/9497 | 0.018348624 | 17.42568807 | 5.599589396 | 0.005531149 | 0.015408201 | 0.008317518 |
| hsa04936 | Human Diseases | Endocrine and metabolic disease | Alcoholic liver disease | 2/10 | 144/9497 | 0.013888889 | 13.19027778 | 4.785477553 | 0.009486706 | 0.024665435 | 0.013314675 |
| hsa04630 | Environmental Information Processing | Signal transduction | JAK-STAT signaling pathway | 2/10 | 168/9497 | 0.011904762 | 11.30595238 | 4.375534661 | 0.012752051 | 0.031083123 | 0.016779014 |
| hsa04621 | Organismal Systems | Immune system | NOD-like receptor signaling pathway | 2/10 | 189/9497 | 0.010582011 | 10.04973545 | 4.079852866 | 0.015960392 | 0.036615016 | 0.019765191 |
| hsa05167 | Human Diseases | Infectious disease: viral | Kaposi sarcoma-associated herpesvirus infection | 2/10 | 196/9497 | 0.010204082 | 9.690816327 | 3.991440758 | 0.017100324 | 0.037050701 | 0.020000378 |
| hsa05171 | Human Diseases | Infectious disease: viral | Coronavirus disease - COVID-19 | 2/10 | 238/9497 | 0.008403361 | 7.980672269 | 3.540866787 | 0.024647703 | 0.050592653 | 0.027310474 |
| hsa05144 | Human Diseases | Infectious disease: parasitic | Malaria | 1/10 | 50/9497 | 0.02 | 18.994 | 4.141633486 | 0.051442036 | 0.098902291 | 0.053388551 |
| hsa04151 | Environmental Information Processing | Signal transduction | PI3K-Akt signaling pathway | 2/10 | 362/9497 | 0.005524862 | 5.246961326 | 2.674756295 | 0.05325508 | 0.098902291 | 0.053388551 |

**Table S6**. Reactome Pathway Enrichment Analysis - This table presents the results of Reactome pathway enrichment analysis. The analysis identifies biological pathways significantly enriched among differentially expressed proteins. Key metrics include gene ratios, enrichment factors, and statistical significance values.

| **Reactome ID** | **Description** | **GeneRatio** | **BgRatio** | **RichFactor** | **FoldEnrichment** | **Z-score** | **p-value** | **p.adjust** | **q-value** |
| --- | --- | --- | --- | --- | --- | --- | --- | --- | --- |
| R-HSA-380108 | Chemokine receptors bind chemokines | 6/10 | 59/11214 | 0.101694915 | 114.040678 | 26.00760335 | 3.37506E-12 | 9.11265E-11 | 3.55269E-11 |
| R-HSA-375276 | Peptide ligand-binding receptors | 6/10 | 201/11214 | 0.029850746 | 33.47462687 | 13.87917781 | 6.08992E-09 | 8.22139E-08 | 3.20522E-08 |
| R-HSA-418594 | G alpha (i) signalling events | 6/10 | 318/11214 | 0.018867925 | 21.15849057 | 10.89462744 | 9.46826E-08 | 8.52143E-07 | 3.3222E-07 |
| R-HSA-373076 | Class A/1 (Rhodopsin-like receptors) | 6/10 | 334/11214 | 0.017964072 | 20.14491018 | 10.61173705 | 1.26766E-07 | 8.55671E-07 | 3.33595E-07 |
| R-HSA-500792 | GPCR ligand binding | 6/10 | 468/11214 | 0.012820513 | 14.37692308 | 8.831411051 | 9.31783E-07 | 5.03163E-06 | 1.96165E-06 |
| R-HSA-449147 | Signaling by Interleukins | 5/10 | 460/11214 | 0.010869565 | 12.18913043 | 7.320902068 | 2.41389E-05 | 0.000108625 | 4.2349E-05 |
| R-HSA-6785807 | Interleukin-4 and Interleukin-13 signaling | 3/10 | 108/11214 | 0.027777778 | 31.15 | 9.405777926 | 9.92488E-05 | 0.000382817 | 0.000149246 |
| R-HSA-6783783 | Interleukin-10 signaling | 2/10 | 47/11214 | 0.042553191 | 47.71914894 | 9.58848415 | 0.000757331 | 0.002555993 | 0.000996488 |
| R-HSA-6788467 | IL-6-type cytokine receptor ligand interactions | 1/10 | 17/11214 | 0.058823529 | 65.96470588 | 8.008025717 | 0.015062627 | 0.043044131 | 0.016781338 |
| R-HSA-5676594 | TNF receptor superfamily (TNFSF) members mediating non-canonical NF-kB pathway | 1/10 | 18/11214 | 0.055555556 | 62.3 | 7.775702628 | 0.015942271 | 0.043044131 | 0.016781338 |
| R-HSA-6783589 | Interleukin-6 family signaling | 1/10 | 24/11214 | 0.041666667 | 46.725 | 6.699133813 | 0.021205303 | 0.049025857 | 0.019113395 |
| R-HSA-210991 | Basigin interactions | 1/10 | 25/11214 | 0.04 | 44.856 | 6.55809565 | 0.022080008 | 0.049025857 | 0.019113395 |
| R-HSA-380994 | ATF4 activates genes in response to endoplasmic reticulum stress | 1/10 | 27/11214 | 0.037037037 | 41.53333333 | 6.299582157 | 0.023827307 | 0.049025857 | 0.019113395 |
| R-HSA-5669034 | TNFs bind their physiological receptors | 1/10 | 29/11214 | 0.034482759 | 38.66896552 | 6.067909003 | 0.025571796 | 0.049025857 | 0.019113395 |
| R-HSA-381042 | PERK regulates gene expression | 1/10 | 32/11214 | 0.03125 | 35.04375 | 5.761386343 | 0.02818327 | 0.049025857 | 0.019113395 |
| R-HSA-1592389 | Activation of Matrix Metalloproteinases | 1/10 | 33/11214 | 0.03030303 | 33.98181818 | 5.66846671 | 0.02905236 | 0.049025857 | 0.019113395 |
| R-HSA-1266695 | Interleukin-7 signaling | 1/10 | 36/11214 | 0.027777778 | 31.15 | 5.412910861 | 0.031655435 | 0.050276279 | 0.019600889 |
| R-HSA-1442490 | Collagen degradation | 1/10 | 64/11214 | 0.015625 | 17.521875 | 3.959918488 | 0.055649646 | 0.08347447 | 0.032543653 |

**SUPPLEMENTAL MATERIAL 2:**

***Table S1****. Differential inflammatory protein expression between Clusters 1 and 2. Log₂ fold change (log2FC), t-statistics, p-values, false discovery rate (FDR, Benjamini–Hochberg correction), and −log₁₀(p) values are shown. Proteins meeting both thresholds (FDR < 0.05 and |log₂FC| ≥ 1) are marked as significant.*

| **Protein** | **log2FC** | **Welch’s t** | **P-value** | **FDR** | **negLog10P** | **Significant** |
| --- | --- | --- | --- | --- | --- | --- |
| **MCP4** | **1.62245703** | **11.8009548** | **5.0639E-16** | **4.6588E-14** | **15.2955138** | **Yes** |
| **CXCL6** | **1.4846631** | **10.3125154** | **4.1172E-14** | **1.8939E-12** | **13.3853936** | **Yes** |
| **CXCL5** | **3.18426113** | **9.76934379** | **1.8562E-12** | **5.6923E-11** | **11.7313762** | **Yes** |
| **IL8** | **1.16732495** | **8.85867253** | **5.9613E-12** | **1.3711E-10** | **11.2246607** | **Yes** |
| **IL7** | **1.07997137** | **8.77162243** | **9.9632E-12** | **1.5641E-10** | **11.0015997** | **Yes** |
| **CXCL1** | **1.73495228** | **8.62831032** | **1.9549E-11** | **2.5693E-10** | **10.7088818** | **Yes** |
| **CXCL11** | **1.32497332** | **7.63995951** | **5.9745E-10** | **6.1072E-09** | **9.22370204** | **Yes** |
| **OSM** | **2.03457387** | **7.9527285** | **7.8756E-10** | **7.2456E-09** | **9.10371641** | **Yes** |
| **MMP1** | **1.88841591** | **7.16190465** | **3.897E-09** | **3.2593E-08** | **8.40927236** | **Yes** |
| **TNFSF14** | **1.01165525** | **6.27282862** | **1.0448E-07** | **5.654E-07** | **6.98098679** | **Yes** |
| VEGFA | 0.79586876 | 9.10932163 | 1.0201E-11 | 1.5641E-10 | 10.9913753 | No |
| MCP1 | 0.80640052 | 8.10039153 | 1.0296E-10 | 1.1841E-09 | 9.98732503 | No |
| HGF | 0.71906113 | 6.70427436 | 1.7258E-08 | 1.3231E-07 | 7.76300132 | No |
| CCL4 | 0.87426396 | 6.63820717 | 2.1484E-08 | 1.5204E-07 | 7.66788369 | No |
| LAP_TGF_beta_1 | 0.7315919 | 6.97457185 | 2.5766E-08 | 1.6932E-07 | 7.58895211 | No |
| CD40 | 0.37847676 | 6.49138499 | 3.2244E-08 | 1.9776E-07 | 7.49154978 | No |
| BetaNGF | 0.15075687 | 6.4229248 | 4.2658E-08 | 2.4529E-07 | 7.3699967 | No |
| CCL11 | 0.6605053 | 5.82479758 | 3.6363E-07 | 1.8585E-06 | 6.4393418 | No |
| CSF1 | 0.30567912 | 5.5946875 | 8.5155E-07 | 4.1233E-06 | 6.06978964 | No |
| MCP2 | 0.94065206 | 4.71782064 | 2.0957E-05 | 9.6403E-05 | 4.67866812 | No |
| LIFR | 0.18724717 | 4.29748115 | 7.6606E-05 | 0.00033028 | 4.11573865 | No |
| CCL28 | 0.88765038 | 4.37237123 | 7.898E-05 | 0.00033028 | 4.10248425 | No |
| FGF5 | 0.20991316 | 4.28018995 | 9.6393E-05 | 0.00038557 | 4.01595558 | No |
| IL2 | 0.31612923 | 4.05996065 | 0.00016614 | 0.00063687 | 3.77952239 | No |
| AXIN1 | 0.87560536 | 3.92463465 | 0.00027376 | 0.00100743 | 3.56263229 | No |
| TWEAK | 0.3473425 | 3.88527928 | 0.00030188 | 0.00106818 | 3.52017078 | No |
| TGFalpha | 0.65109459 | 4.02798177 | 0.00034778 | 0.00114272 | 3.45869103 | No |
| FGF23 | 0.39848085 | 3.84195352 | 0.00033869 | 0.00114272 | 3.47019312 | No |
| IL6 | 0.66537577 | 3.79005774 | 0.00050107 | 0.00158961 | 3.30009909 | No |
| MCP3 | 0.7267847 | 3.56390063 | 0.00087053 | 0.00266964 | 3.0602143 | No |
| CXCL10 | 0.56474739 | 3.29553749 | 0.00181002 | 0.00537166 | 2.74231736 | No |
| IL18 | 0.45034981 | 3.19199748 | 0.00251083 | 0.00721865 | 2.60018203 | No |
| CD244 | 0.38222725 | 3.04128775 | 0.00430929 | 0.01177884 | 2.36559469 | No |
| TSLP | 0.44179071 | 2.9820517 | 0.00435305 | 0.01177884 | 2.36120624 | No |
| CD8A | 0.40727777 | 2.95644299 | 0.00468573 | 0.01197465 | 2.32922263 | No |
| CCL3 | 0.58003874 | 3.01023253 | 0.00463335 | 0.01197465 | 2.33410446 | No |
| IL10RB | 0.22937753 | 2.92268739 | 0.00513938 | 0.012779 | 2.28908918 | No |
| IL20 | 0.26767492 | 2.96739644 | 0.00538362 | 0.01303403 | 2.26892552 | No |
| LIF | 0.45684736 | 2.94447094 | 0.00554268 | 0.01307504 | 2.25628017 | No |
| SCF | 0.2560839 | 2.86339675 | 0.00626671 | 0.01406189 | 2.20296026 | No |
| IL15RA | 0.21398549 | 2.85707692 | 0.00624672 | 0.01406189 | 2.204348 | No |
| CX3CL1 | 0.25930467 | 2.70380015 | 0.00943074 | 0.02065781 | 2.02545429 | No |
| TNF | 0.32097132 | 2.64104313 | 0.01110024 | 0.02374935 | 1.9546676 | No |
| OPG | 0.20219635 | 2.58566133 | 0.01282976 | 0.02682586 | 1.89178142 | No |
| IL24 | 0.51616451 | 2.52233776 | 0.01655235 | 0.03384036 | 1.78114037 | No |
| CCL23 | 0.28418264 | 2.41699114 | 0.01921881 | 0.03843762 | 1.71627354 | No |
| CCL25 | 0.31237016 | 2.38025012 | 0.02100899 | 0.04112397 | 1.67759492 | No |
| PDL1 | 0.24700615 | 2.24434464 | 0.02926184 | 0.0560852 | 1.53369835 | No |
| CXCL9 | 0.34765467 | 2.15224695 | 0.0365743 | 0.06794211 | 1.43682398 | No |
| IL10 | 0.28845382 | 2.14380487 | 0.03692506 | 0.06794211 | 1.4326788 | No |
| IL13 | -0.209523 | -2.0179745 | 0.05018665 | 0.09053277 | 1.29941182 | No |
| FGF21 | 0.69206808 | 1.91201311 | 0.06164143 | 0.10905792 | 1.21012727 | No |
| ARTN | 0.25103168 | 1.90535453 | 0.06285128 | 0.10910033 | 1.2016859 | No |
| TNFB | 0.26447846 | 1.89328199 | 0.06505792 | 0.11083941 | 1.18669985 | No |
| IL22RA1 | 0.36874838 | 1.83833152 | 0.07179225 | 0.12008885 | 1.14392245 | No |
| TNFRSF9 | 0.15054766 | 1.80756572 | 0.0770854 | 0.1266403 | 1.11302787 | No |
| ST1A1 | 0.51608514 | 1.76418689 | 0.08490655 | 0.13704216 | 1.07105878 | No |
| ENRAGE | 0.48332918 | 1.63889722 | 0.10733062 | 0.17024856 | 0.96927638 | No |
| CD5 | 0.13097813 | 1.60259035 | 0.1151057 | 0.17692435 | 0.93890318 | No |
| DNER | 0.11703228 | 1.60123179 | 0.11538544 | 0.17692435 | 0.93784898 | No |
| CST5 | 0.21866937 | 1.57891383 | 0.12045871 | 0.18167543 | 0.91916179 | No |
| SLAMF1 | 0.14080396 | 1.55580891 | 0.12589204 | 0.18680755 | 0.90000172 | No |
| CASP8 | 0.29678404 | 1.54479664 | 0.12886259 | 0.18818029 | 0.88987315 | No |
| IL2RB | 0.14173981 | 1.42331167 | 0.16108615 | 0.23156135 | 0.79294179 | No |
| IL33 | 0.14325019 | 1.40819906 | 0.1654665 | 0.23419874 | 0.78128991 | No |
| CDCP1 | 0.19470022 | 1.39163693 | 0.1711456 | 0.23856659 | 0.76663426 | No |
| IL17C | 0.26826775 | 1.35828864 | 0.18033822 | 0.2476286 | 0.74391223 | No |
| GDNF | 0.26576385 | 1.34034435 | 0.18816982 | 0.25458269 | 0.72545004 | No |
| CCL19 | -0.2611943 | -1.3301976 | 0.19206595 | 0.25608794 | 0.71654962 | No |
| CCL20 | 0.31484203 | 1.31105296 | 0.19567584 | 0.25717396 | 0.70846279 | No |
| Flt3L | 0.12100423 | 1.26920897 | 0.2100702 | 0.27220364 | 0.67763556 | No |
| TRAIL | 0.10361879 | 1.20690134 | 0.23388507 | 0.29885315 | 0.6309975 | No |
| MMP10 | 0.19843277 | 1.06970647 | 0.29026749 | 0.36537421 | 0.5372016 | No |
| STAMBP | 0.38922135 | 1.06049327 | 0.29388795 | 0.36537421 | 0.53181822 | No |
| NT3 | 0.18215407 | 0.96575663 | 0.3386376 | 0.41539546 | 0.47026482 | No |
| IL12B | 0.12438008 | 0.93818187 | 0.3526866 | 0.42693641 | 0.45261104 | No |
| X4EBP1 | 0.4453519 | 0.8609896 | 0.39328861 | 0.46990327 | 0.40528863 | No |
| NRTN | 0.17649157 | 0.84930049 | 0.39976256 | 0.47151481 | 0.39819788 | No |
| IL4 | -0.1196149 | -0.8406157 | 0.40530293 | 0.47199835 | 0.39222025 | No |
| IFNgamma | 0.21160107 | 0.79099878 | 0.43312321 | 0.49809169 | 0.36338854 | No |
| IL10RA | 0.14897665 | 0.75811684 | 0.45180459 | 0.5072877 | 0.34504936 | No |
| SIRT2 | 0.38127637 | 0.75766049 | 0.45214773 | 0.5072877 | 0.34471964 | No |
| IL20RA | -0.1034946 | -0.7039539 | 0.48661771 | 0.53938349 | 0.31281209 | No |
| IL1alpha | 0.08059563 | 0.64695249 | 0.52051413 | 0.5700869 | 0.28356748 | No |
| TRANCE | -0.0722001 | -0.3711834 | 0.71230634 | 0.77096687 | 0.14733319 | No |
| IL18R1 | 0.03845277 | 0.33541954 | 0.73878028 | 0.79032309 | 0.1314847 | No |
| CD6 | 0.03922107 | 0.31572324 | 0.75347654 | 0.79677979 | 0.12293026 | No |
| IL5 | -0.0665603 | -0.2570105 | 0.7982658 | 0.83455061 | 0.09785248 | No |
| IL17A | 0.05758302 | 0.23093774 | 0.8184289 | 0.84601639 | 0.08701904 | No |
| uPA | 0.01343049 | 0.18698556 | 0.85239968 | 0.86673509 | 0.06935672 | No |
| FGF19 | 0.04971769 | 0.18068911 | 0.85731406 | 0.86673509 | 0.06686006 | No |
| ADA | -0.0187937 | -0.1172451 | 0.90713743 | 0.90713743 | 0.04232691 | No |
